# COVID-19 pandemic: examining the faces of spatial differences in the morbidity and mortality in sub-Saharan Africa, Europe and USA

**DOI:** 10.1101/2020.04.20.20072322

**Authors:** Adebayo A. Otitoloju, Ifeoma P. Okafor, Mayowa Fasona, Kafilat A. Bawa-Allah, Chukwuemeka Isanbor, Onyeka S. Chukwudozie, Olawale S. Folarin, Taiwo O. Adubi, Temitope O. Sogbanmu, Anthony E. Ogbeibu

## Abstract

**Background:** COVID-19, the disease associated with the Severe Acute Respiratory Syndrome Coronavirus-2 (SARS-CoV-2) is currently a global pandemic with several thousands of confirmed cases of infection and death. However, the death rate across affected countries shows variation deserving of critical evaluation.

**Methods:** In this study, we evaluated differentials in COVID-19 confirmed cases of infection and associated deaths of selected countries in Sub-Sahara Africa (Nigeria and Ghana), South Africa, Europe (Italy, Spain, Sweden and UK) and USA. Data acquired for various standard databases on mutational shift of the SARS-CoV-2 virus based on geographical location, BCG vaccination policy, malaria endemicity, climatic conditions (temperature), differential healthcare approaches were evaluated over a period of 45 days from the date of reporting the index case.

**Results:** The number of confirmed cases of infection and associated deaths in Sub-Sahara Africa were found to be very low compared to the very high values in Europe and USA over the same period. Recovery rate from COVID-19 is not correlated with the mutational attributes of the virus with the sequenced strain from Nigeria having no significant difference (p>0.05) from other geographical regions. Significantly higher (p<0.05) infection rate and mortality from COVID-19 were observed in countries (Europe and USA) without a current universal BCG vaccination policy compared to those with one (Sub-Sahara African countries). Countries with high malaria burden had significantly lower (p<0.05) cases of COVID-19 than those with low malaria burden. A strong negative correlation (−0.595) between mean annual temperature and COVID-19 infection and death was observed with 14.8% variances between temperature and COVID-19 occurrence among the countries. A clear distinction was observed in the COVID-19 disease management between the developed countries (Europe and USA) and Sub-Sahara Africa.

**Conclusions:** The study established that the wide variation in the outcome of the COVID-19 disease burden in the selected countries are attributable largely to climatic condition (temperature) and differential healthcare approaches to management of the disease. We recommend consideration and mainstreaming of these findings for urgent intervention and management of COVID-19 across these continents.

## 1.0 Introduction

Coronavirus disease 2019 (COVID-19) is the disease associated with Severe Acute Respiratory Syndrome Coronavirus-2 (SARS-CoV-2). Characterising COVID-19 as a pandemic is an acknowledgement that the coronavirus disease 2019 (COVID-19) outbreak, which started in the Hubei province of China in 2019, has now spread to all continents, affecting most countries around the world including African countries (Cucinotta and Vanelli, 2020). Since the outbreak of COVID-19, it has spread through different countries and continents, but with differential impacts and peculiarities. The European region has recorded about 1.05 million confirmed cases and 93,000 deaths; Regions of Americas – 743,000 confirmed cases and 33,000 deaths; Western Pacific Region – 127,000 confirmed cases and 5558 deaths; Eastern Mediterranean Region – 115,000 confirmed cases and 5,600 deaths; South-east Asia region – 23,000 confirmed cases and 1000 deaths; and Africa 12,000 confirmed cases and 500 deaths (WHO, 2020).

It has been anticipated that the impacts of Covid-19 outbreak in African countries and many developing countries would be very devastating because of several reasons including poor state of health infrastructure, poverty concerns, unfavourable living conditions in the cities, population densities and prevalence of underlying disease conditions like lower respiratory infection, malaria, diarrheal, HIV/AIDS and tuberculosis (The World Bank, 2020). In addition, many West African countries have poorly resourced health systems, rendering them unable to quickly scale up responses to disease outbreaks (Baker, 2020). Most countries in the region have fewer than five hospital beds per 10,000 of the population and fewer than two medical doctors per 10,000 of the population. In contrast to Italy and Spain with 34 and 35 hospital beds respectively per 10,000 of the population and about 41 medical doctors per 10,000 of the population (Martinez-Alvarez, 2020). According to UNECA (2020), Africa could see 300,000 deaths from the coronavirus this year even under the best-case scenario. Under the worst-case scenario with no interventions against the virus, Africa could see 3.3 million deaths and 1.2 billion infections. The fears being expressed by many of these genuine observers of potential dangers that may occur in Africa as a result of the COVID-19 epidemic are therefore well-founded because all the metrics point to a very catastrophic outcome as it is currently being recorded in some countries in Latin America with similar level of development and poor healthcare system (AFP News Agency – www.aljazeera.com).

Despite all the concerns of poor healthcare systems and underlying poor conditions, the outbreak of COVID-19 in some parts of Africa has not resulted in the largely anticipated huge spike in the number of confirmed cases and deaths. This surprisingly low number after a thirty (30) to forty-five (45) day period of the COVID-19 outbreak in some of the African countries is raising a lot of research questions in order to understand the true situation of the epidemic in these countries and also provide lessons learnt or to be learnt around the world (Ebenso and Otu, 2020; Vaughan, 2020).

It is therefore pertinent to investigate the trend in the spread of this disease, establish similarities or disparities among countries and attempt to determine the underlying reasons for the observed variations. Furthermore, there is an urgent demand to ensure that adequate preventive measures are put in place to ensure that the disease does not spread among very vulnerable populations where the healthcare system is at best weak and may not be capable of coping with the epidemic, unlike the more robust healthcare systems in Europe and America (Gilbert *et. al*., 2020).

The main objectives of this paper are to compare COVID-19 confirmed cases and deaths data in some countries in Africa (Nigeria, Ghana and South Africa) with those reported in the more developed countries (Italy, Spain, UK, Sweden and USA) with better healthcare systems. There is therefore the need to evaluate several underlying factors in the spatial differences observed with incidence and disease burden of COVID-19. Specific factors examined in this paper are: mutational shift of the SARS-CoV-2 virus based on geographical location, BCG vaccination policy of the countries, malaria endemicity, climatic conditions (temperature), differential healthcare approaches as dictated by cultural norms, practice and unfettered access to prescription drugs.

## 2.0 Methods

### 2.1 Study Countries

The geographical location and basic climatic conditions of the eight study countries are as follow:

i. Nigeria (Lon. 3°-14°E; Lat. 4°-14°N) and Ghana (Lon.1°E-3°W; Lat. 5°-11°N) are located in the low latitude tropics with no marked summer-winter differentials.
ii. Republic of South Africa (Lon.17°-33°E; Lat. 23°-35°S) is located immediately outside the tropic of Capricorn in the southern hemisphere. It experiences an inverted boreal climate with a rainy winter (May to September) and hot and dry summer (October to April).
iii. Italy (Lon. 7°-19°E; Lat. 37°-46°N), Spain (Lon. 11°W-4°E; Lat. 36°-44°N), United Kingdom (Lon. 8°W-2°E; Lat. 50°-58°N), Sweden (Lon. 12°-24°E; Lat. 55°-68°N) and the Mainland United States (Lon. 67°-125°W; Lat. 25°-50°N) are located in the northern mid-latitudes between the tropic of Cancer and the Arctic Circle. They experience distinct summer (May to August) and winter (October to March).

### 2.2 Data Collection Sources

Data on confirmed cases of COVID-19 infection and associated reported deaths were gathered from the eight study countries over a period of 46 days starting from first index case. Cumulative COVID-19 infection and death records were sourced from the COVID-19 dashboard of the Center for Systems Science and Engineering of John Hopkins University (https://coronavirus.jhu.edu/map.html) and Worldometer (https://www.worldodometers.info). The data were cross checked with data of National Centres for Disease Control for the relevant countries for accuracy.

The deposited curated genomic data of the sequenced virus (SARS-Cov-2) from different geographical locations was retrieved from the National Center for Biotechnology Information (NCBI). Their respective accession number and number of base pairs are summarised in Table 1. The sequenced data from Nigeria is deposited in (https://github.com/acegid/CoV_Sequences).

**Table 1:** Retrieved SARS-CoV-2 genomic data

| Country | Genbank ID | No of base pairs |
| --- | --- | --- |
| China | MN908947 | 29903 bp |
| *Nigeria | Not deposited | 29417 bp |
| Japan | LC528232.1 | 29902 bp |
| California, USA | MN985325.1 | 29882 bp |
| Australia | MT007544.1 | 29893 bp |
| India | MT012098.1 | 29854 bp |
| South Korea | MT039890.1 | 29903 bp |
| Finland | MT020781.2 | 29806 bp |
| Italy | MT066156.1 | 29867 bp |
| Nepal | MT072688.1 | 29811 bp |
| Sweden | MT093571.1 | 29886 bp |
| Brazil | MT126808.1 | 29876 bp |
| Spain | MT233519.1 | 29782 bp |
| Pakistan | MT262993.1 | 29836 bp |
| Peru | MT263074.1 | 29856 bp |
| Israel | MT276597.1 | 29851 bp |
| France | MT320538.1 | 29679 bp |
| Iran | MT320891.1 | 29828 bp |
| South Africa | MT324062.1 | 29903 bp |
| Turkey | MT327745.1 | 29832 bp |
| Greece | MT328032.1 | 29903 bp |

\*Nigeria data source: (https://github.com/acegid/CoV_Sequences)

Data on Bacille Calmette-Guerin (BCG) vaccination policy for the selected countries was collected from www.bcgatlas.org.

Recent data on malaria cases in these countries (Nigeria, Ghana and South Africa) were retrieved from World Malaria Report (WHO, 2019). For USA data was retrieved from CDC website (www.cdc.gov) while for Spain, Italy, Sweden and UK from Europe annual epidemiological report (ECDC, 2019).

Mean Temperature data for 50 years (1962 to 2012) for all the study countries was sourced from the archive of the Stat World (https://stat.world/biportal/?allsol=1). The temperature data were summarized for long-term spatial and temporal means.

Information on healthcare approaches were obtained from anecdotal data, reviews of literature and statistic websites like www.nairametrics.com and www.healthdata.org.

### 2.4 Data Analysis

#### 2.4.1 Statistical Analysis of COVID-19 Confirmed Cases of Infection and Deaths

Data on confirmed cases of Coronavirus infection and reported deaths from complications were gathered from the 8 study countries over a period of 46 days starting from first index case.

These data were analysed for significant relationships between time, confirmed cases of infection and deaths using simple linear correlation and regression models with the purpose of developing predictive equations to establish future trend and projection in each country. Multivariate statistics using similarity indices and hierarchical cluster analysis were performed to identify similarities or disparities among countries in terms of the chosen parameters -confirmed cases and reported deaths (Hammer et al., 2001; Ogbeibu, 2014).

Mann-Whitney test and Spearman’s rank correlation were done to determine relationship between BCG vaccination, malaria endemicity and climatic condition (temperature) with level of occurrence of COVID-19 confirmed cases and deaths in study countries. Level of significance is set at 5% (p<0.05).

#### 2.4.2 Evaluation of possible mutational shift of novel Coronavirus (SARS-CoV-2) based on geographical location

The SARS-CoV-2 sequences from different geographical locations were properly edited using a bio-edit tool software, as gaps within the sequence were deleted. A sequence alignment analysis was conducted using the Clustal W, which is a multiple sequence alignment program that uses seeded guide trees and HMM profile-profile techniques to generate alignments between three or more sequences, then utilizing the UPGMA/Neighbor-joining method to generate a distance matrix. The program also uses the BLOSUM scoring matrix, as default settings of gap penalty at 5, gap open penalty at 15 and finally, gap penalty cost at 6.66 per element.

##### 2.4.2.1 Phylogenetic Analysis

An automated tree was generated after the consistent alignments. The tree was rendered using the RaxML bootstrap (Stamatakis, 2006). The resulting clustering shows the relationships between the countries.

## 3.0 Results

### 3.1 Outcome of Statistical Analysis of COVID-19 Confirmed Cases of Infection and Deaths

#### 3.1.1 Daily Trend in Confirmed Cases of infection and Deaths

The results of the daily trend of confirmed cases of infections and deaths over a 45 day period for Nigeria, Ghana, South Africa, Italy, Spain, UK, USA and Sweden are presented in Figures 1 & 2. The results indicate differential upward trend in the number of confirmed cases with USA having the highest number of cases followed by Italy, Spain, UK, Sweden, Ghana and Nigeria (least) over the 45 days period (Fig. 1). For the recorded number of deaths from COVID-19, Italy has the highest number of deaths from the disease followed by Spain, USA, UK, Sweden, Ghana and Nigeria (least) over the 45 days period (Fig. 2).

**Fig. 1:**
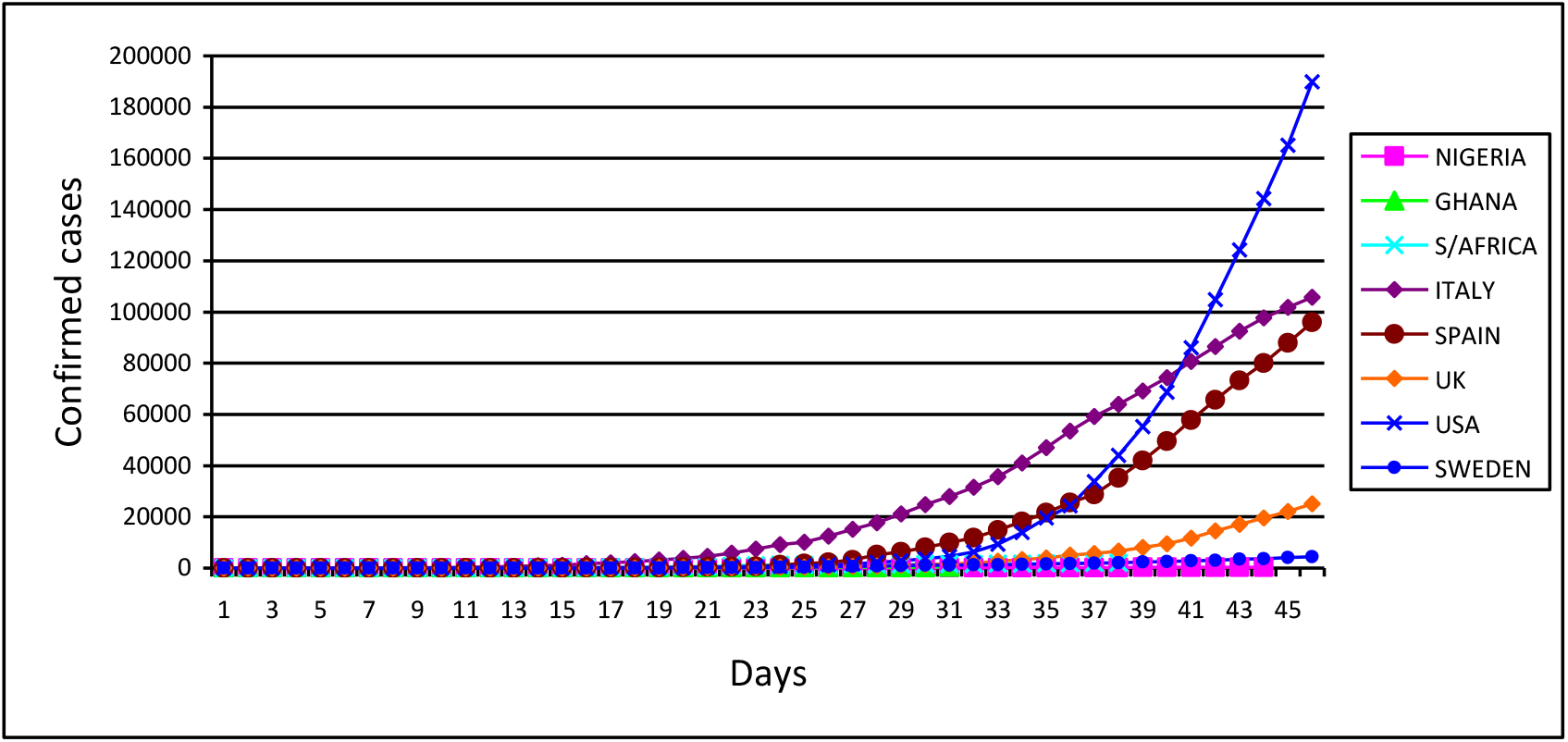
Daily Confirmed cases of coronavirus infection in eight countries

**Fig. 2:**
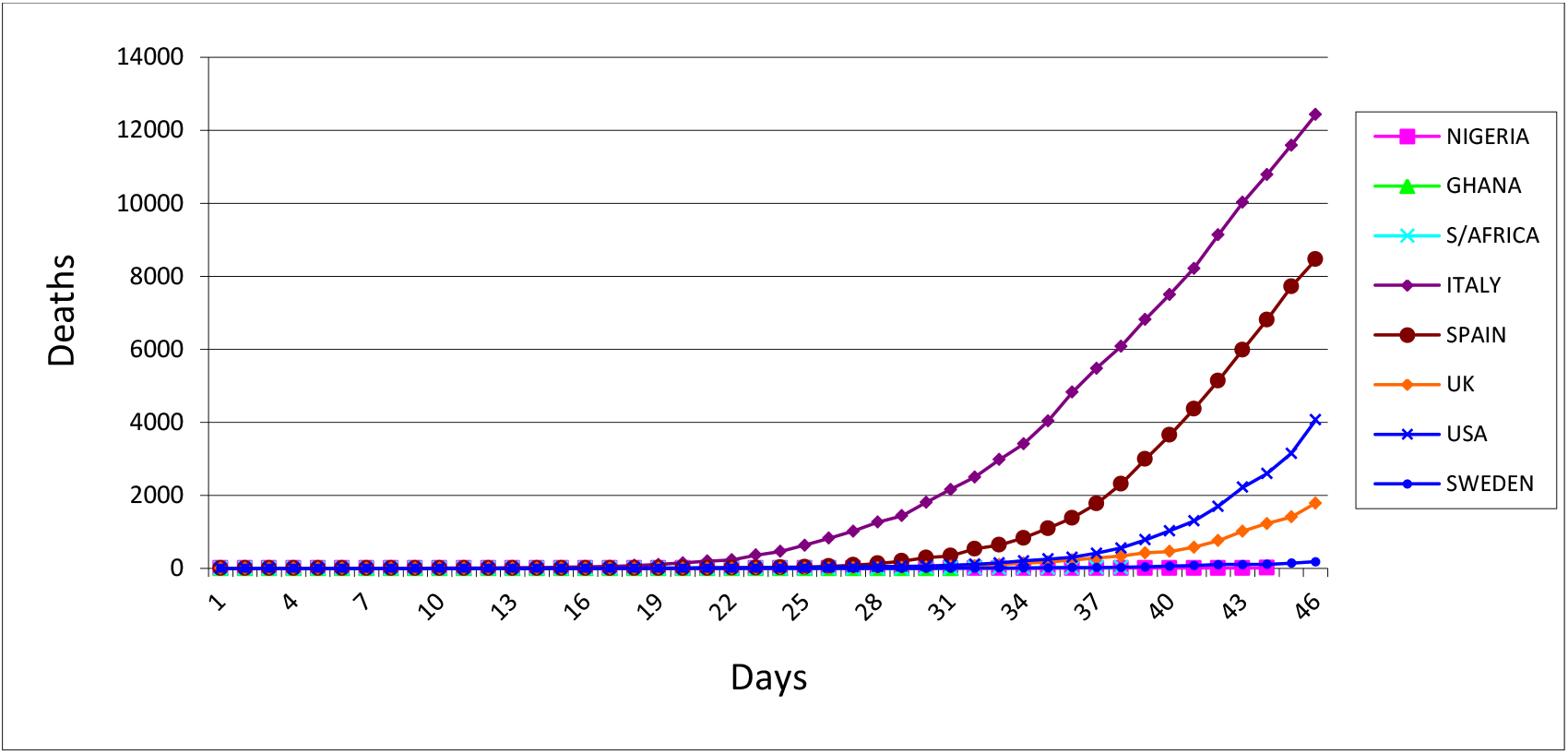
Daily Deaths from COVID-19 infection in eight countries

#### 3.1.2 Projections on confirmed cases of infection over time in different countries

In order to understand and monitor the daily trend in cases of infection and death, using available data, regression and correlation analysis were performed to establish relationships and generate predictive regression equations for each country. The higher the coefficient of determination R^2^, the higher the proportion of confirmed cases of infection and deaths accounted for by time. On the basis of the analysis, a severity classification index was derived to establish the time – infection (confirmed cases) (Table 2) and time – death relationships for the countries (Table 3).

**Table 2:** Time - Infection relationships and Regression Equations for different countries

| Country | Significance of Regression<br>(P-Value) | Regression Equation<br>$Y = bX \pm a$ | Coefficient of determination<br>$R^2$ | Remark on projected rate of increase |
| --- | --- | --- | --- | --- |
| Nigeria | 7.43E-16 | $y = 7.2376x - 82.165$ | 0.7907 | Very low |
| Ghana | 9.95E-17 | $y = 13.223x - 78.439$ | 0.9104 | Very low |
| South Africa | 1.99E-21 | $y = 64.424x - 491.95$ | 0.921 | Low |
| Italy | 1.03E-16 | $y = 2244.2x - 26353$ | 0.7944 | Very high |
| Spain | 4.26E-11 | $y = 1592.1x - 21142$ | 0.6317 | Very high |
| UK | 1.68E-09 | $y = 359.88x - 4877.2$ | 0.5657 | High |
| USA | 2.79E-08 | $y = 2544.6x - 35672$ | 0.5077 | Very high |
| Sweden | 6.24E-16 | $y = 83.126x - 993.52$ | 0.7769 | Low |

*Severity Classification based on 'b': < 50 Very Low, 51-100 Low, 101-500 High, >500 Very high*

**Table 3:**
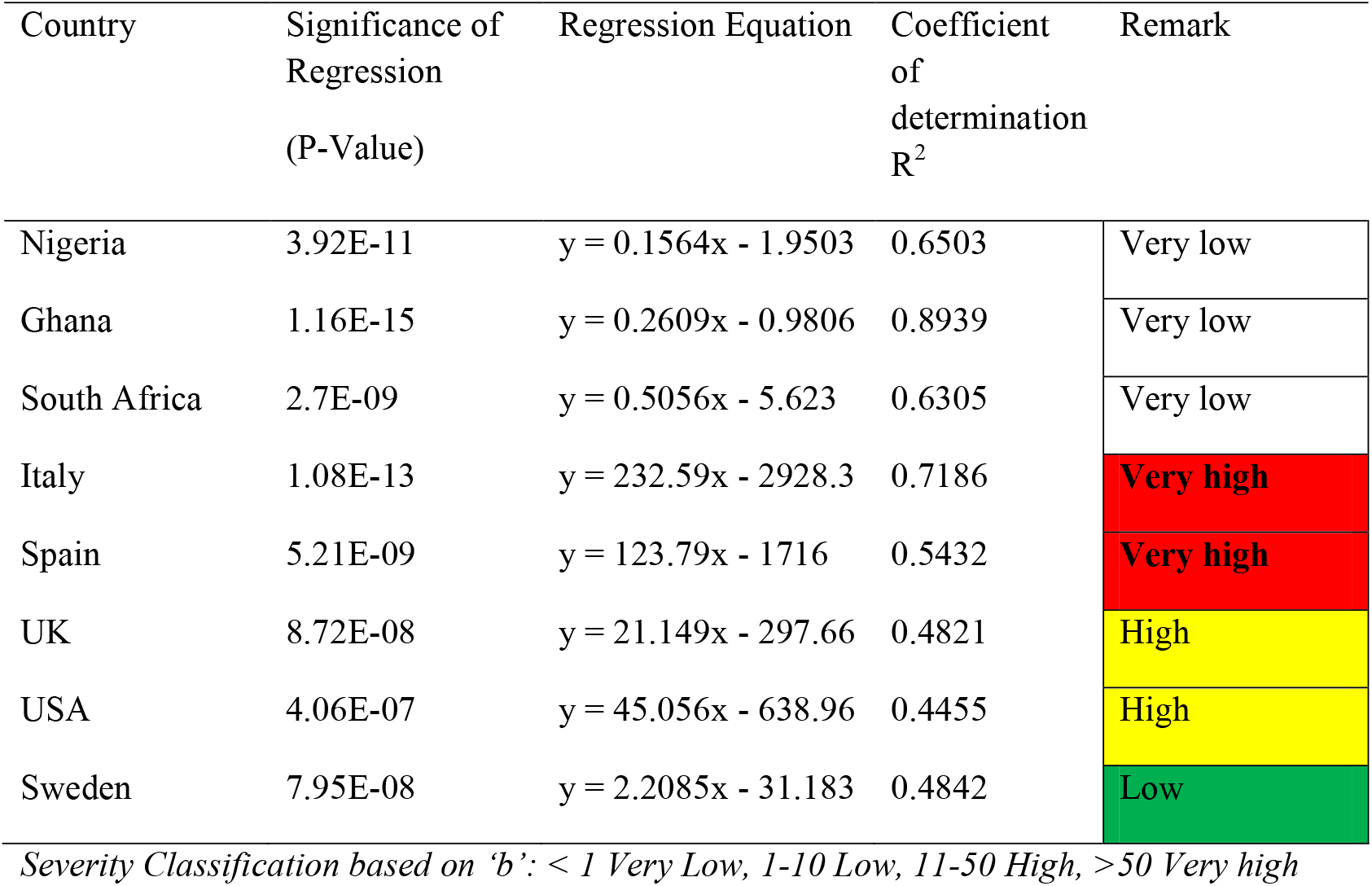
Time - Death relationships and Regression Equations for different countries

| Country | Significance of Regression<br>(P-Value) | Regression Equation | Coefficient of determination<br>$R^2$ | Remark |
| --- | --- | --- | --- | --- |
| Nigeria | 3.92E-11 | $y = 0.1564x - 1.9503$ | 0.6503 | Very low |
| Ghana | 1.16E-15 | $y = 0.2609x - 0.9806$ | 0.8939 | Very low |
| South Africa | 2.7E-09 | $y = 0.5056x - 5.623$ | 0.6305 | Very low |
| Italy | 1.08E-13 | $y = 232.59x - 2928.3$ | 0.7186 | Very high |
| Spain | 5.21E-09 | $y = 123.79x - 1716$ | 0.5432 | Very high |
| UK | 8.72E-08 | $y = 21.149x - 297.66$ | 0.4821 | High |
| USA | 4.06E-07 | $y = 45.056x - 638.96$ | 0.4455 | High |
| Sweden | 7.95E-08 | $y = 2.2085x - 31.183$ | 0.4842 | Low |

*Severity Classification based on 'b': < 1 Very Low, 1-10 Low, 11-50 High, >50 Very high*

For the number of confirmed cases, Nigeria and Ghana were projected to be **VERY LOW**, South Africa and Sweden as **LOW**, United Kingdom (UK) as **HIGH**, while USA, Italy and Spain were projected to be **VERY HIGH** (Table 2).

For the number of deaths, Nigeria, Ghana and South Africa were projected to be **VERY LOW**, Sweden as **LOW**, United Kingdom (UK) and USA as **HIGH**, while Italy and Spain were projected to be **VERY HIGH** (Table 3).

#### 3.1.3 Relationship between Confirmed Cases and Reported Deaths in different countries

In order to establish the relationship between confirmed cases and death numbers, using available data, regression and correlation analysis were performed to establish the relationships in reported values. On the basis of the analysis, a classification index was derived and used to classify relationship of confirmed cases of COVID-19 infection and deaths over time for the countries. On the basis of the derived indices, the relationship for Nigeria, Ghana, South Africa, USA and Sweden were classified as LOW while time-death relationships for Italy, Spain and UK were classifies as HIGH (Table 4).

In order to group the countries on the basis of similarity in trends of confirmed cases and deaths, Similarity and distance indices (Bray Curtis and Euclidean) were computed among countries. These indices were further supported with hierarchical cluster analysis.

**Table 4:**
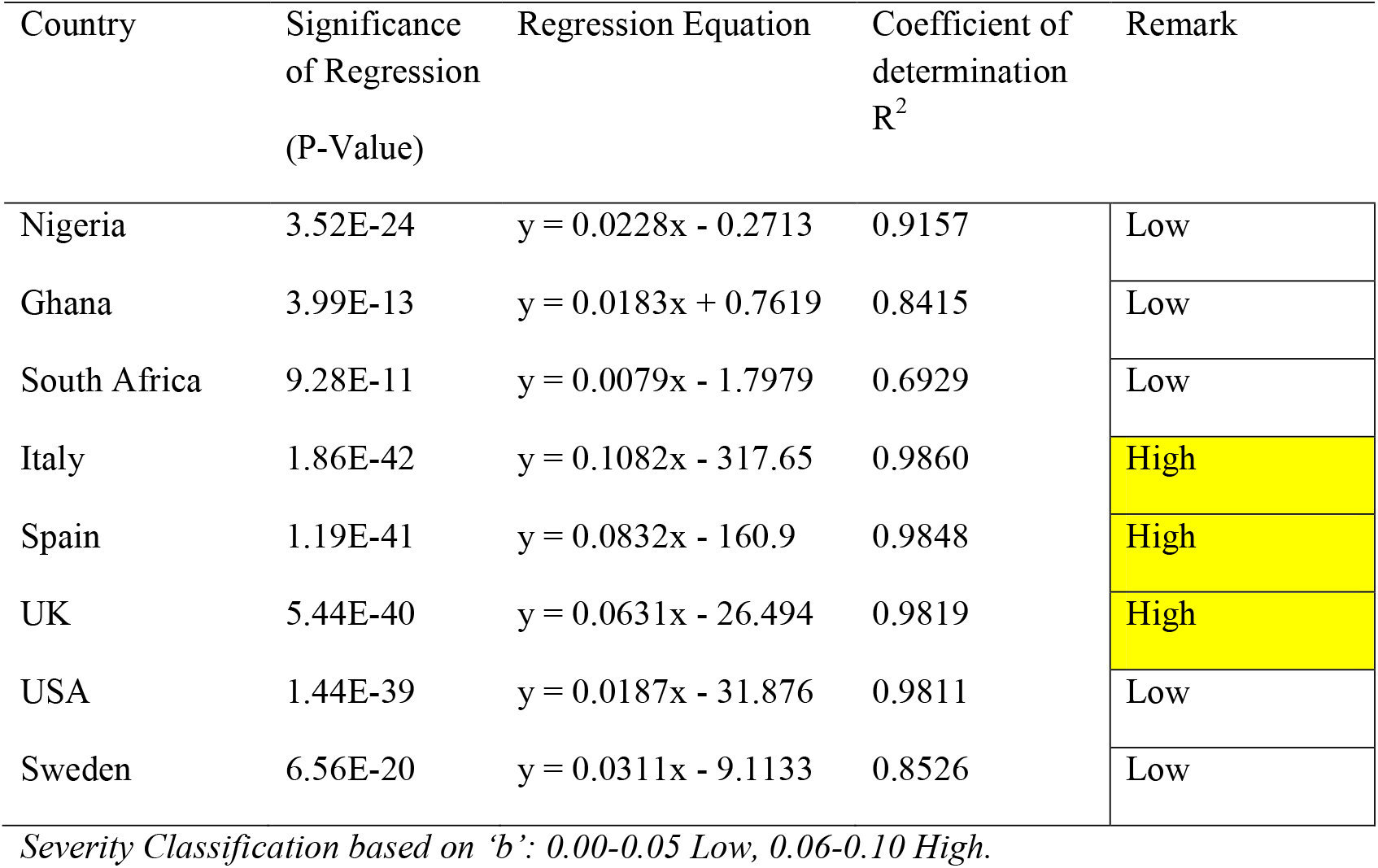
Confirmed Infection - Death relationships and Regression Equations for different countries

The results of the similarity and distance indices for the confirmed cases of COVID-19 infection are presented in Tables 5 - 6 and Figures 3 - 4. The dendrogram clustering divided the countries into 2 broad groups of Nigeria and Ghana versus the other countries. Further grouping of the other countries was also established with USA, Italy and Spain in a sub-group, South Africa and Sweden in another sub-group while UK was a standalone, although more associated with SA-Sweden sub-group (Figs. 3 - 4).

**Table 5:**
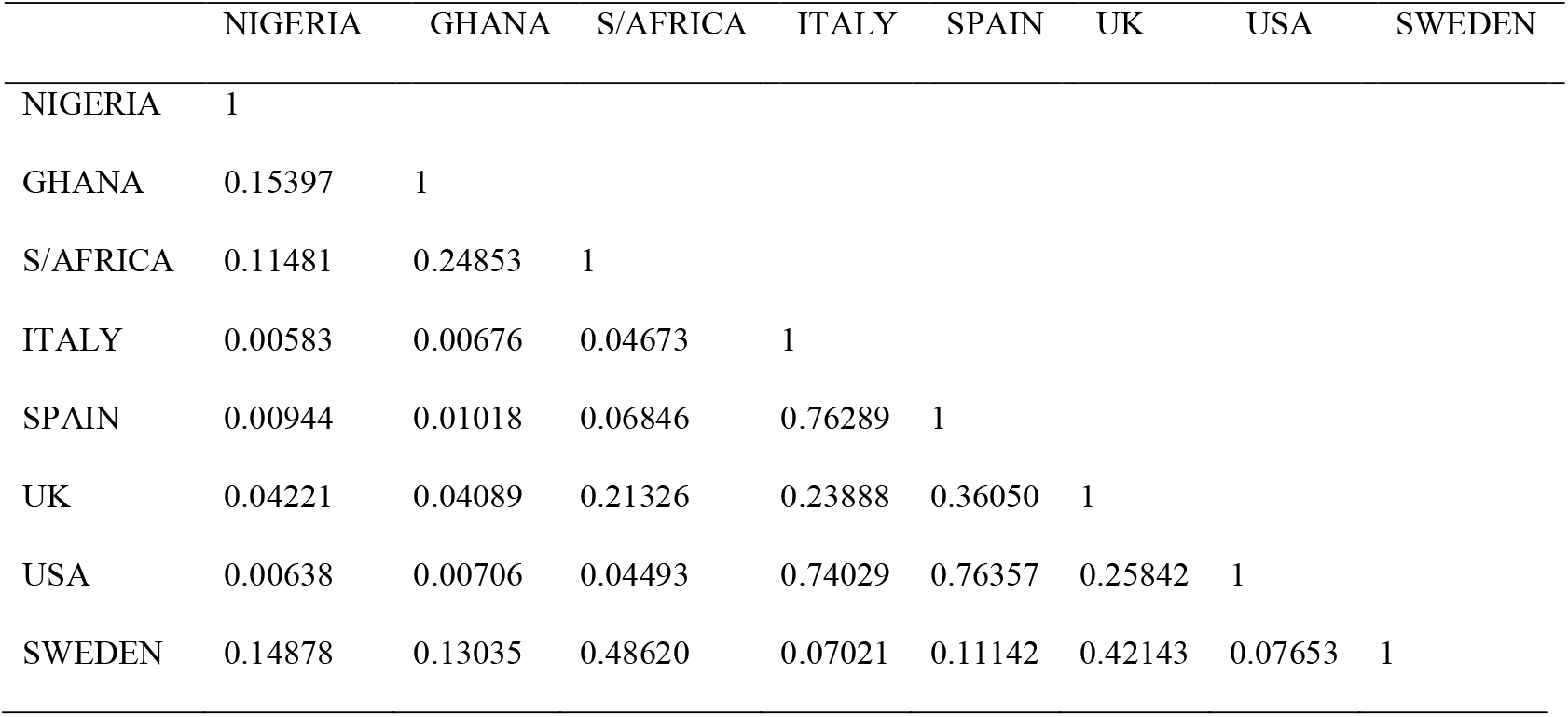
Bray Curtis similarity and distance indices based on Confirmed Cases of Infection

|  | NIGERIA | GHANA | S/AFRICA | ITALY | SPAIN | UK | USA | SWEDEN |
| --- | --- | --- | --- | --- | --- | --- | --- | --- |
| NIGERIA | 1 |  |  |  |  |  |  |  |
| GHANA | 0.15397 | 1 |  |  |  |  |  |  |
| S/AFRICA | 0.11481 | 0.24853 | 1 |  |  |  |  |  |
| ITALY | 0.00583 | 0.00676 | 0.04673 | 1 |  |  |  |  |
| SPAIN | 0.00944 | 0.01018 | 0.06846 | 0.76289 | 1 |  |  |  |
| UK | 0.04221 | 0.04089 | 0.21326 | 0.23888 | 0.36050 | 1 |  |  |
| USA | 0.00638 | 0.00706 | 0.04493 | 0.74029 | 0.76357 | 0.25842 | 1 |  |
| SWEDEN | 0.14878 | 0.13035 | 0.48620 | 0.07021 | 0.11142 | 0.42143 | 0.07653 | 1 |

**Table 6:** Euclidean Similarity and distance indices for Confirmed Cases of Coronavirus infection

|  | NIGERIA | GHANA | S/AFRICA | ITALY | SPAIN | UK | USA | SWEDEN |
| --- | --- | --- | --- | --- | --- | --- | --- | --- |
| NIGERIA | 0 |  |  |  |  |  |  |  |
| GHANA | 1192.5 | 0 |  |  |  |  |  |  |
| S/AFRICA | 6099 | 5943.3 | 0 |  |  |  |  |  |
| ITALY | 2.88E+05 | 2.89E+05 | 2.86E+05 | 0 |  |  |  |  |
| SPAIN | 2.11E+05 | 2.11E+05 | 2.10E+05 | 96436 | 0 |  |  |  |
| UK | 48861 | 49413 | 48549 | 2.43E+05 | 1.62E+05 | 0 |  |  |
| USA | 3.60E+05 | 3.61E+05 | 3.60E+05 | 1.48E+05 | 1.57E+05 | 3.12E+05 | 0 |  |
| SWEDEN | 9996.3 | 10557 | 9867.3 | 2.78E+05 | 2.01E+05 | 39278 | 3.51E+05 | 0 |

**Fig. 3:**
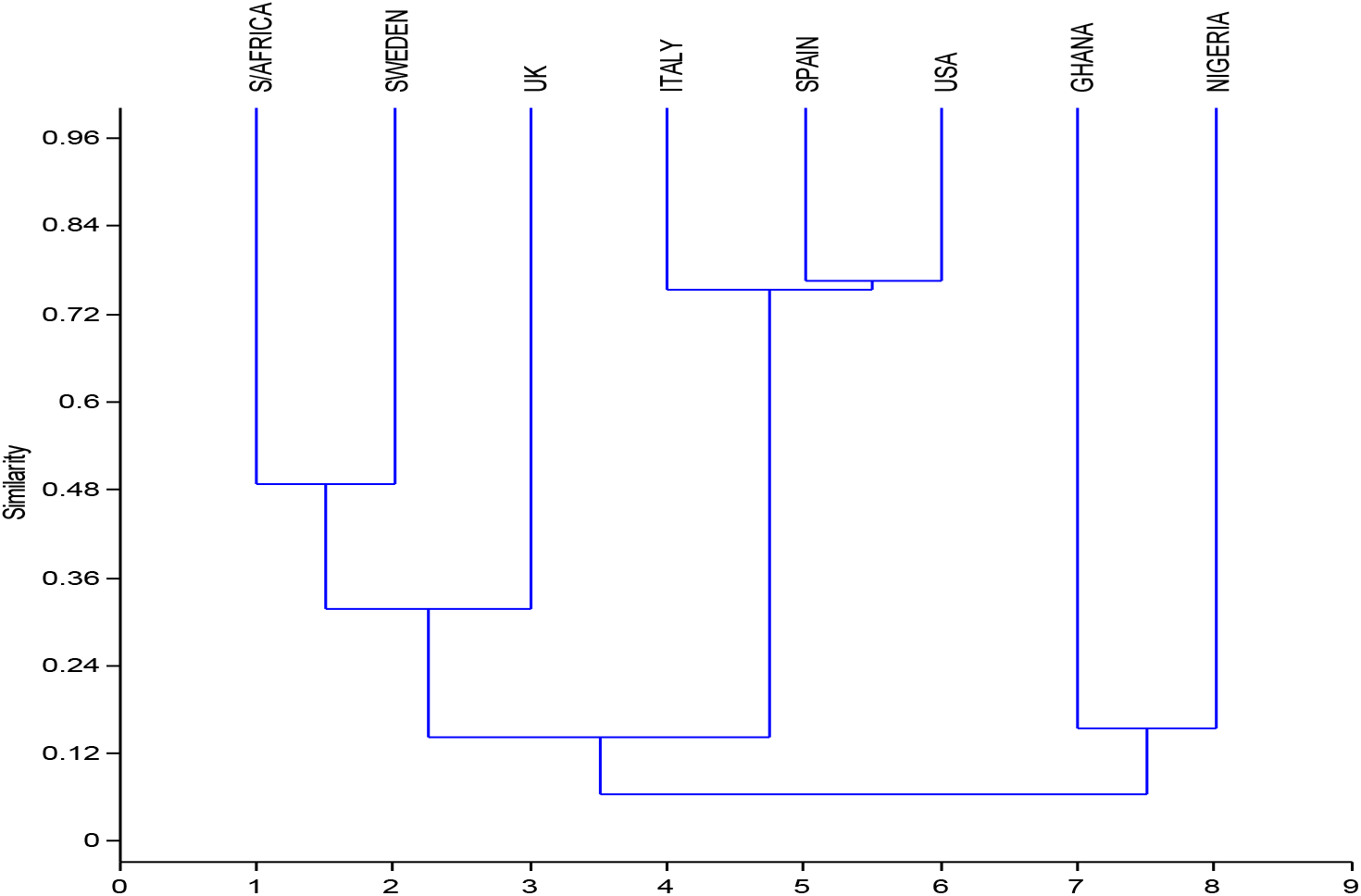
Hierarchical Cluster Analysis of Similarities among Countries based on Confirmed Cases of Coronavirus infection, using Bray-Curtis

**Fig. 4:**
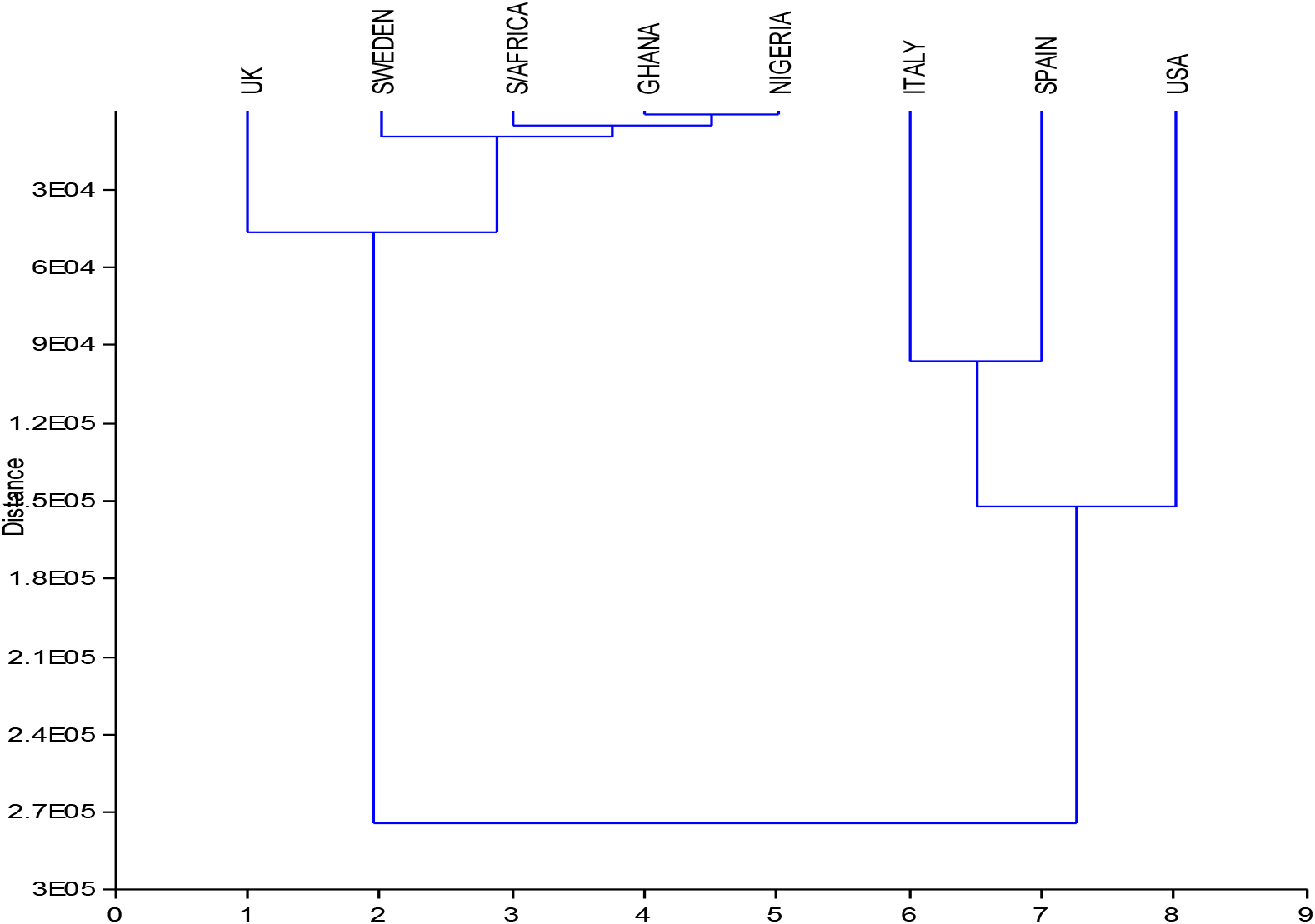
Hierarchical Cluster Analysis of Similarities among Countries based on Confirmed Cases of Coronavirus infection using Euclidean Distance

#### 3.1.4 Similarity and Hierarchical Cluster Analysis

The results of the similarity and distance indices for the number of death cases are presented in Tables 7 - 8 and Figures 5 - 6. The dendrogram clustering divided the countries into 2 broad groups of Nigeria, Ghana, South Africa and Sweden in one broad group while Italy, Spain, UK and USA are clustered in a another broad group. Further sub-groupings of the countries were established (Figures 5 - 6).

**Table 7:** Bray Curtis similarity and distance indices based on Deaths

|  | NIGERIA | GHANA | S/AFRICA | ITALY | SPAIN | UK | USA | SWEDEN |
| --- | --- | --- | --- | --- | --- | --- | --- | --- |
| NIGERIA | 1 |  |  |  |  |  |  |  |
| GHANA | 0.08333 | 1 |  |  |  |  |  |  |
| S/AFRICA | 0.24348 | 0.27692 | 1 |  |  |  |  |  |
| ITALY | 0.00118 | 0.00170 | 0.002755 | 1 |  |  |  |  |
| SPAIN | 0.00251 | 0.00244 | 0.00585 | 0.63959 | 1 |  |  |  |
| UK | 0.01494 | 0.01165 | 0.034509 | 0.14568 | 0.28635 | 1 |  |  |
| USA | 0.00712 | 0.00855 | 0.016534 | 0.28395 | 0.51917 | 0.64387 | 1 |  |
| SWEDEN | 0.13307 | 0.02852 | 0.22262 | 0.016197 | 0.03414 | 0.18828 | 0.094045 | 1 |

**Table 8:**
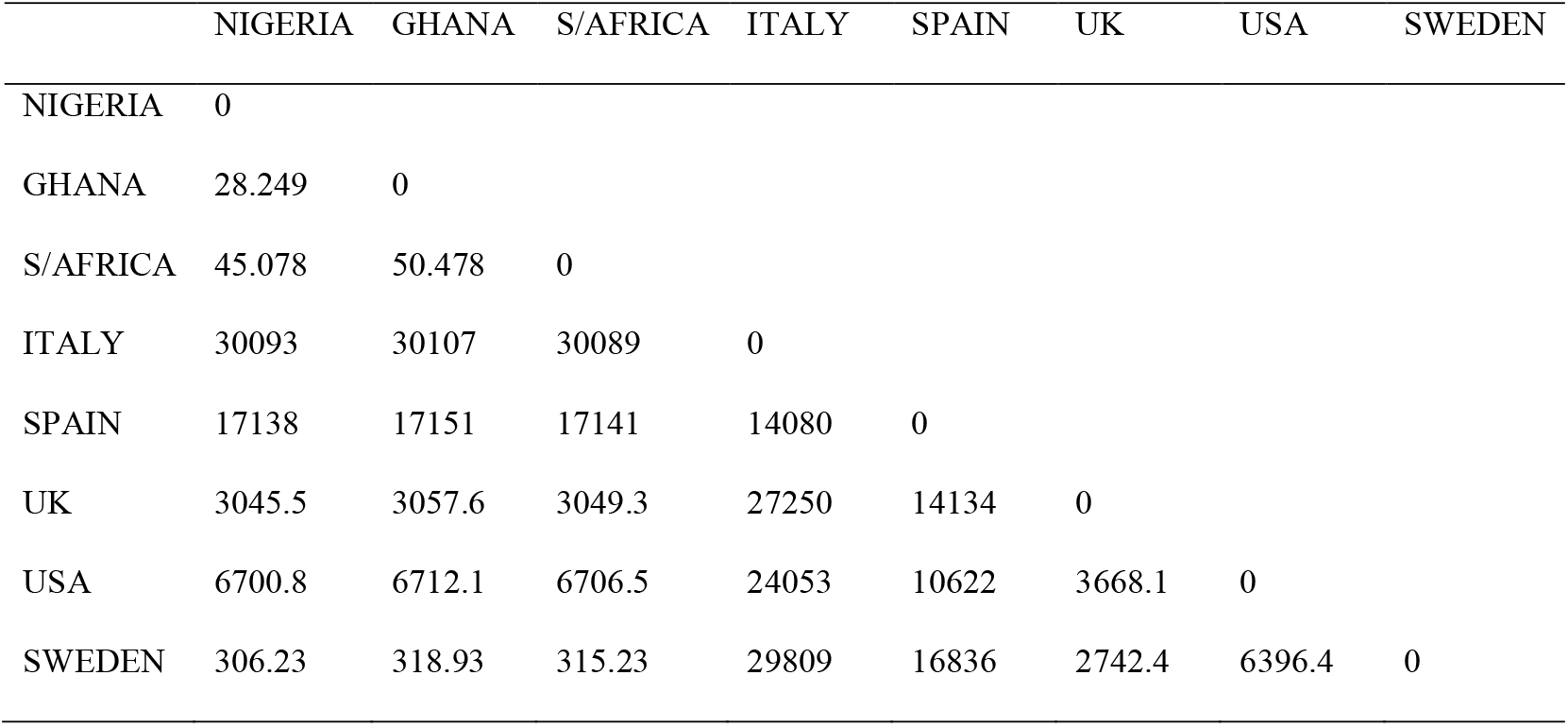
Euclidean similarity and distance indices based on Deaths from infection

**Fig. 5:**
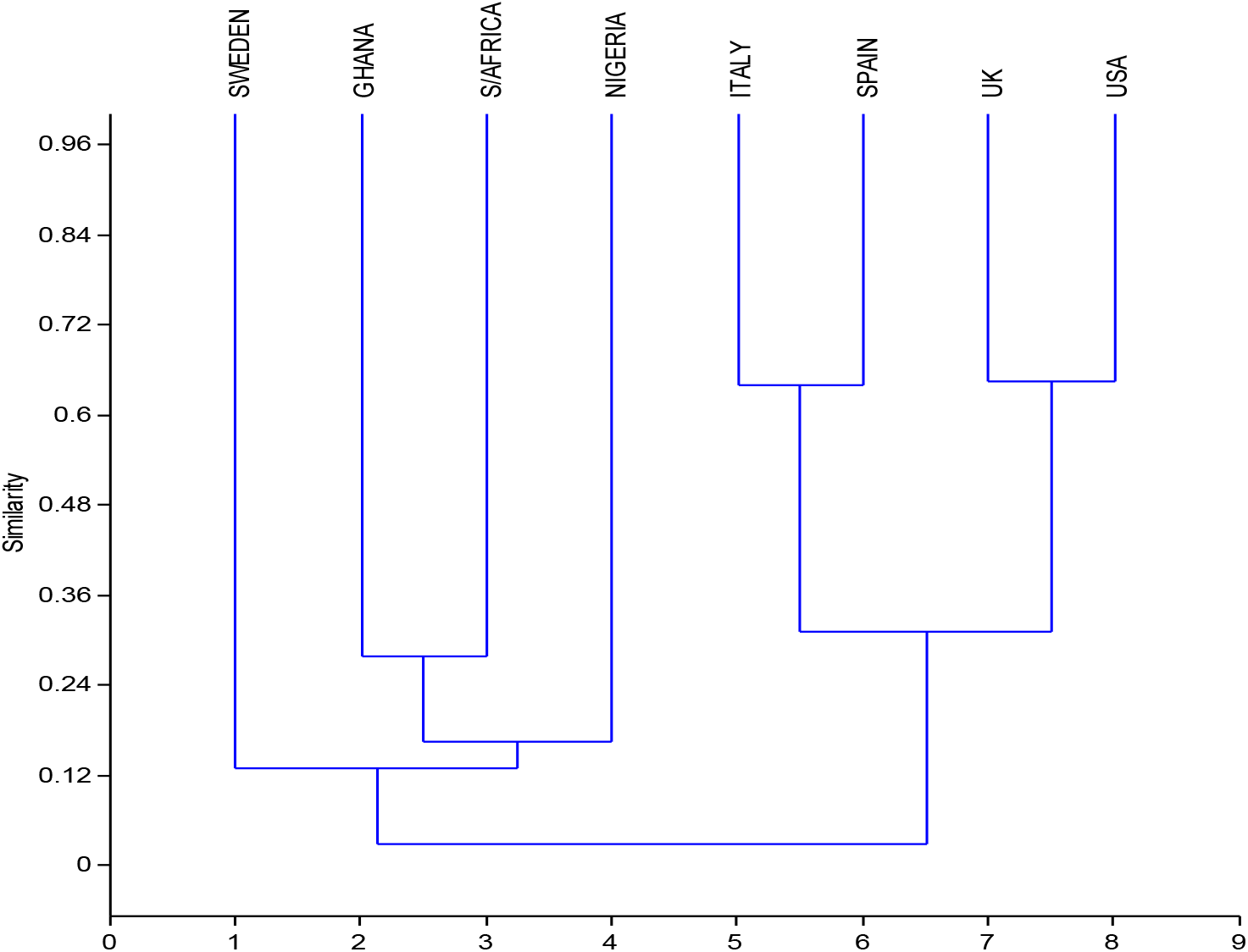
Hierarchical Cluster Analysis of Similarities among Countries based on Deaths from Coronavirus infection using Bray-Curtis

**Fig. 6:**
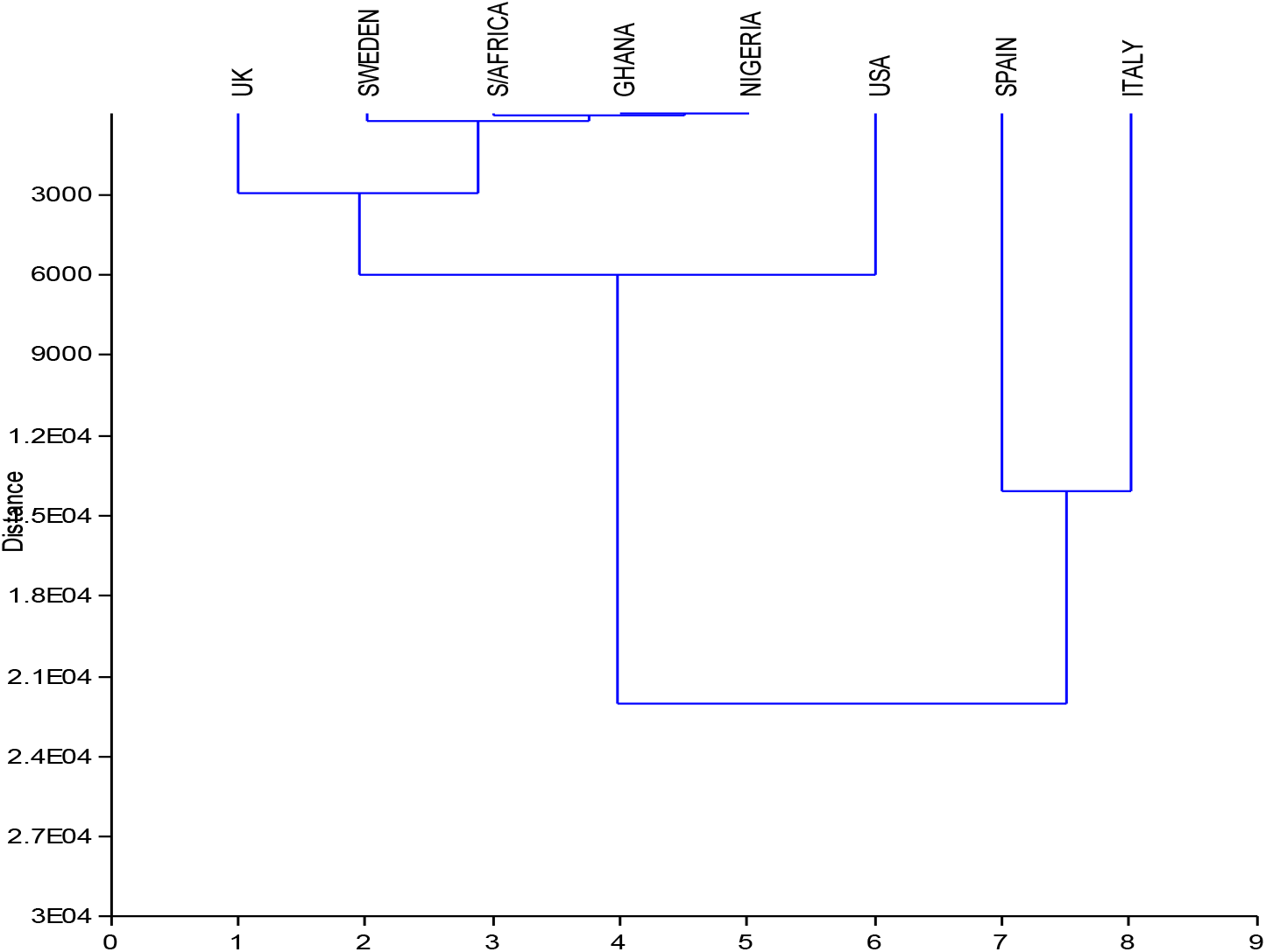
Hierarchical Cluster Analysis of Similarities among Countries based on Deaths from Coronavirus infection using Euclidean Distance

### 3.2 Evaluation of possible mutational shift of novel Coronavirus (SARS-CoV-2) based on geographical location

The number of base pairs is approximately the same for all of the retrieved genome. They were within the range of 29417 bp-29903 bp. The retrieved sequence from the Nigerian population had the least genomic content of 29417 bp, while the South African population had the highest genomic content of 29902 bp. The multiple sequence alignment showed that majority of the genomic regions were highly conserved with little or no polymorphic attributes across the entire genome (Fig. 7). This was further validated from the obtained phylogenetic tree bootstrap value, which revealed that the mutational rate is less significant, with little or no change across the population groups. The bootstrap scores were relatively zero, showing no significant change in the viral genome. The conservation scores for the entire sequence alignment was 98%.

**Fig. 7:**
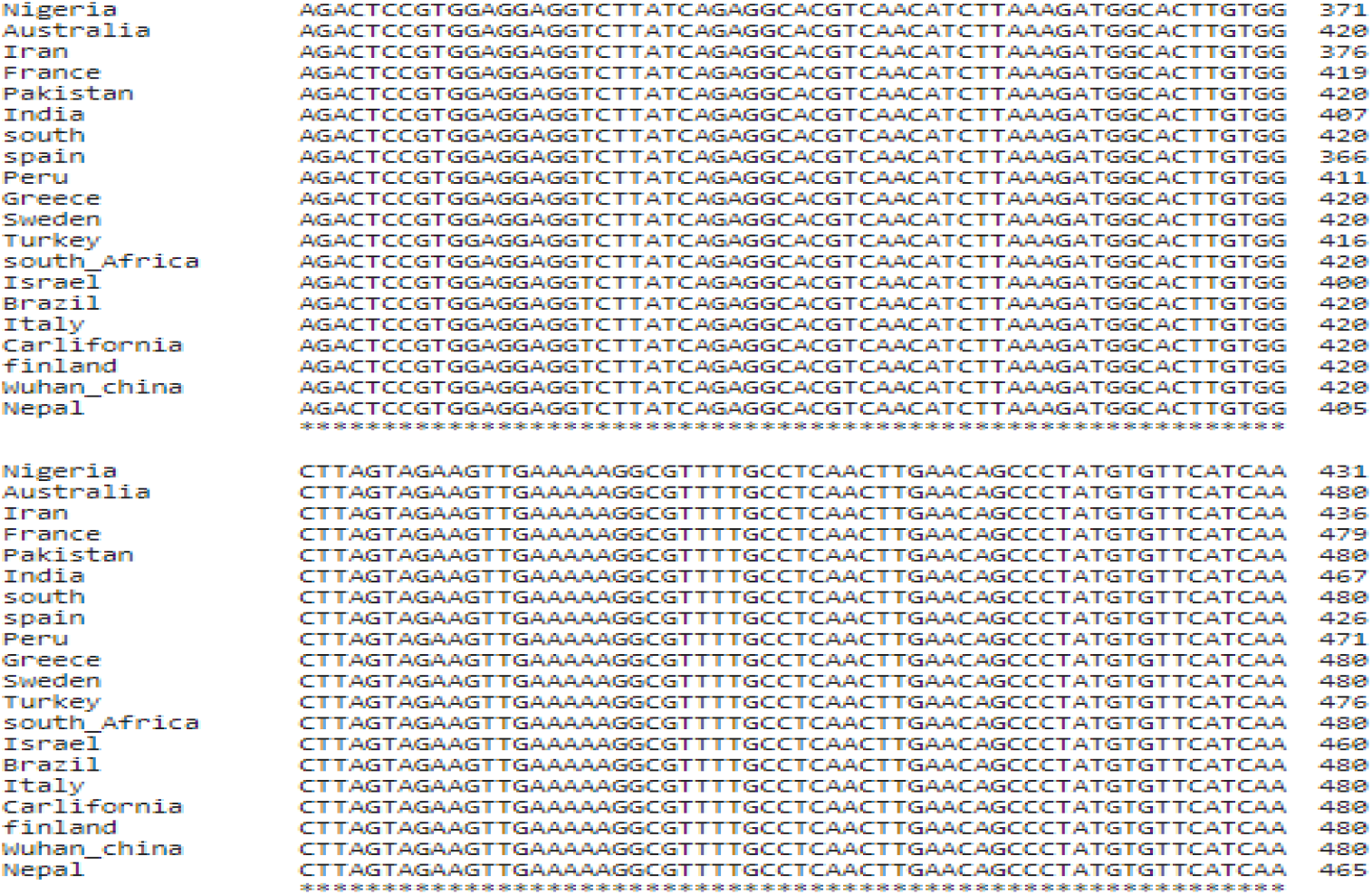
Multiple sequence alignment of the SARS-CoV-2 part genome from different geographical locations. NB: * conserved region

For the phylogenetic analysis, an automated tree was generated after the consistent alignments. The resulting clustering based on phylogenetic analysis shows the relationships between the countries (Fig. 8).

**Fig. 8:**
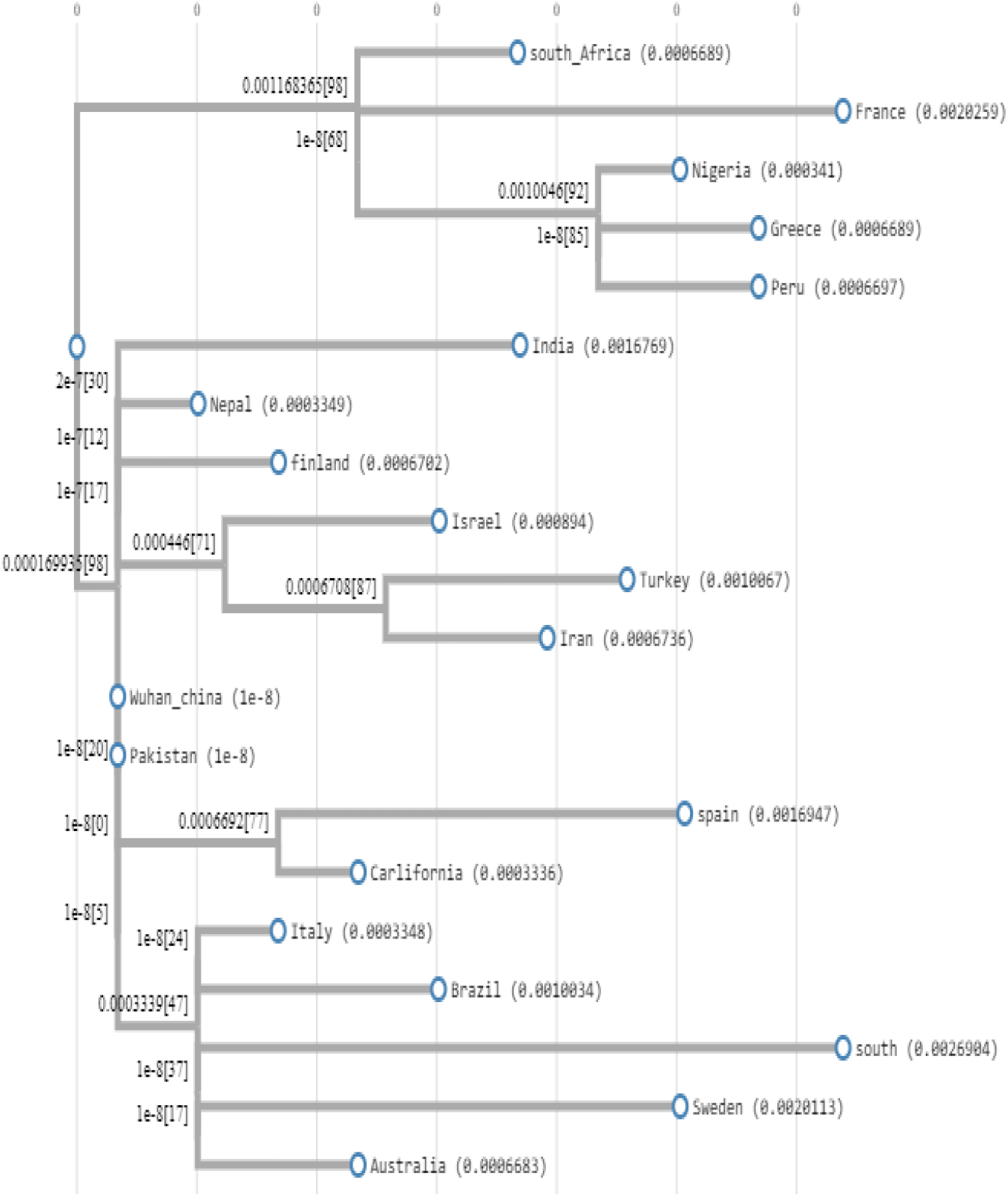
Phylogenetic Tree based on alignment of the concatenated nucleotide sequences of the SARS-Cov-2 virus from different geographical locations. The bootstrap values are found on each node. Nigeria is clustered with the viral strain from Peru and Greece.

### 3.3 Evaluation of the relevance of Bacille Calmette-Guerin (BCG) vaccination policy on COVID-19 incidence and mortality rates

Information collected on BCG vaccination policy in the selected African countries (Nigeria, Ghana and South Africa) showed that all the countries have a current universal BCG vaccination policy, which is administered at birth. Nigeria has a multiple BCG vaccination policy while South Africa has a single vaccination policy. All the developed countries assessed do not have a current universal BCG vaccination policy. Spain, Sweden and the United Kingdom had a universal BCG vaccination policy in the past, but this has been suspended in these countries. The policy was suspended in 1975 in Sweden, in 1981 in Spain and in 2005 in the United Kingdom. Italy and USA have never had a universal BCG vaccination policy (Table 9).

**Table 9:** Bacille Calmette-Guerin (BCG) vaccination policy and incidence of COVID-19 in selected African and Developed countries respectively

| Countries | <sup>a</sup> Total Population (as at 14<sup>th</sup> April 2020) | <sup>b</sup>Current BCG vaccination policy | <sup>b</sup>BCG vaccination policy in the past? | <sup>b</sup>Time of BCG vaccination | <sup>b</sup>Multiple BCG? | <sup>b</sup>Multiple BCG in the past? | Date of first case of SARS-CoV-2 infection | <sup>a</sup>Incidence of SARS-CoV-2 infection (as at 13<sup>th</sup> April) | <sup>a</sup>Death from COVID-19 (as at 13<sup>th</sup> April) | % of the total population infected | % of death from infected population |
| --- | --- | --- | --- | --- | --- | --- | --- | --- | --- | --- | --- |
| Nigeria | 205,042,002 | Yes (policy for all) | Yes | At birth | Yes | Yes | 27<sup>th</sup> February | 343 | 10 | 0.00017 | 2.9 |
| Ghana | 30,934,252 | Yes (policy for all) | Yes | At birth | - | - | 12<sup>th</sup> march | 566 | 8 | 0.00182 | 1.4 |
| South Africa | 59,150,335 | Yes (policy for all) | Yes | At birth | No | Yes | 5<sup>th</sup> March | 2,272 | 27 | 0.00384 | 1.19 |
| Italy | 60,480,342 | No | No | - | No | No | 31<sup>st</sup> January | 159,516 | 20,465 | 0.26 | 12.83 |
| Spain | 46,750,998 | No | Yes (policy for all, stopped 1981) | At birth | No | No | 31<sup>st</sup> January | 170,099 | 17,756 | 0.36 | 10.44 |
| Sweden | 10,086,030 | No | Yes (policy for all, stopped 1975) | At birth | No | Yes | 31<sup>st</sup> January | 10,948 | 919 | 0.11 | 8.39 |
| United Kingdom | 67,811,150 | No | Yes (policy for all, stopped 2005) | 12/13 yrs | No | No | 30<sup>th</sup> January | 88,621 | 11,329 | 0.13 | 12.78 |
| USA | 330,594,891 | No | No | - | No | No | 20<sup>th</sup> January | 586,941 | 23,641 | 0.18 | 4.03 |

<sup>a</sup> source: www.worldometers.info
<sup>b</sup> source: www.bcgatlas.org
SARS-CoV-2: severe acute respiratory syndrome coronavirus 2 (virus causing COVID-19)
COVID-19: Coronavirus disease 2019

Analysis of the data BCG policy in selected countries and confirmed cases of COVID-19 showed that there is a significantly (p<0.05) higher infection rate in countries that do not have a current universal BCG vaccination policy compared to those countries with a vaccination policy (Figure 9). The percentage death due to COVID-19 is also significantly (p < 0.05) higher in countries without the BCG vaccination policy compared to countries with existing BCG policy (Figure 10). However, there was no significant (p > 0.05) difference in infections rates in countries which had a universal vaccination policy in the past compared to countries that never had a universal vaccination policy (Figure 11).

**Fig. 9.**
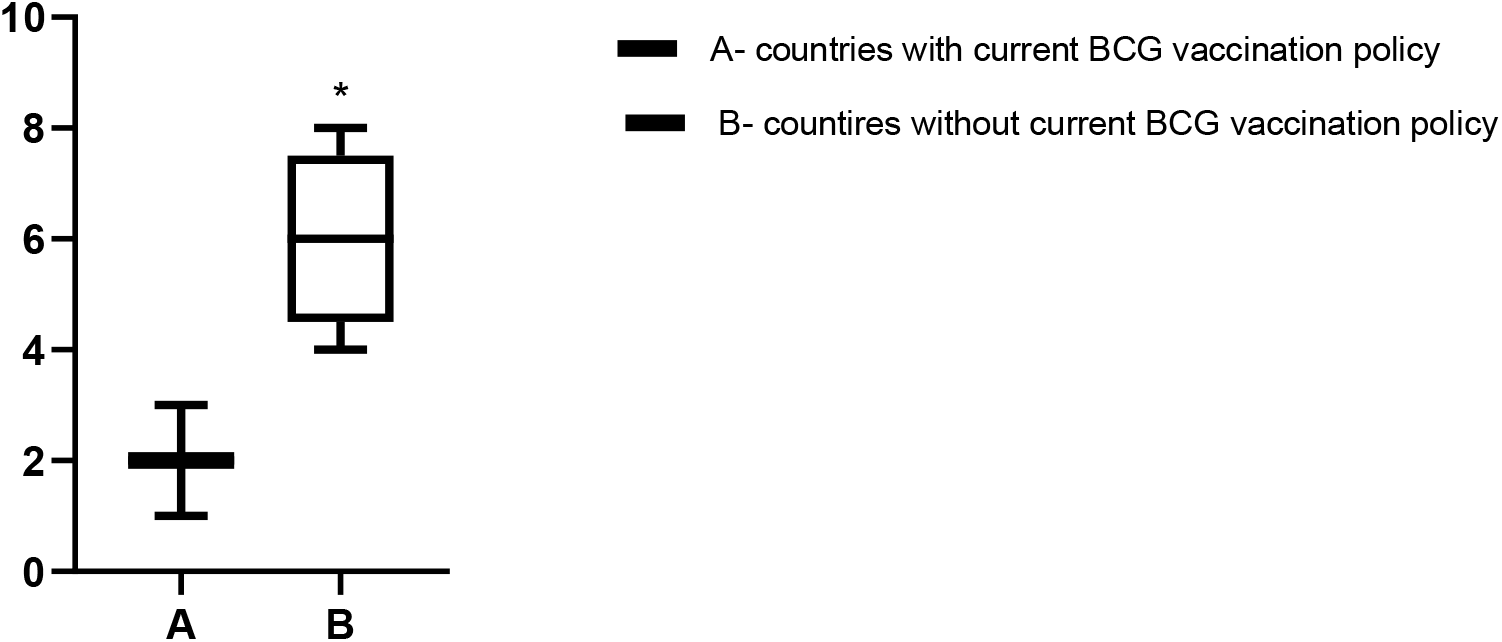
Percentage of the total population infected with SARS-CoV-2 in **A**- countries with a current universal BCG vaccination policy (Nigeria, Ghana and South Africa) compared to **B**- countries without a current universal BCG vaccination policy (Italy, Spain, Sweden, UK and USA). Rank plot using Mann Whitney non-parametric test shows a significant (p < 0.05) difference in infection rates in these two groups of countries

**Fig. 10.**
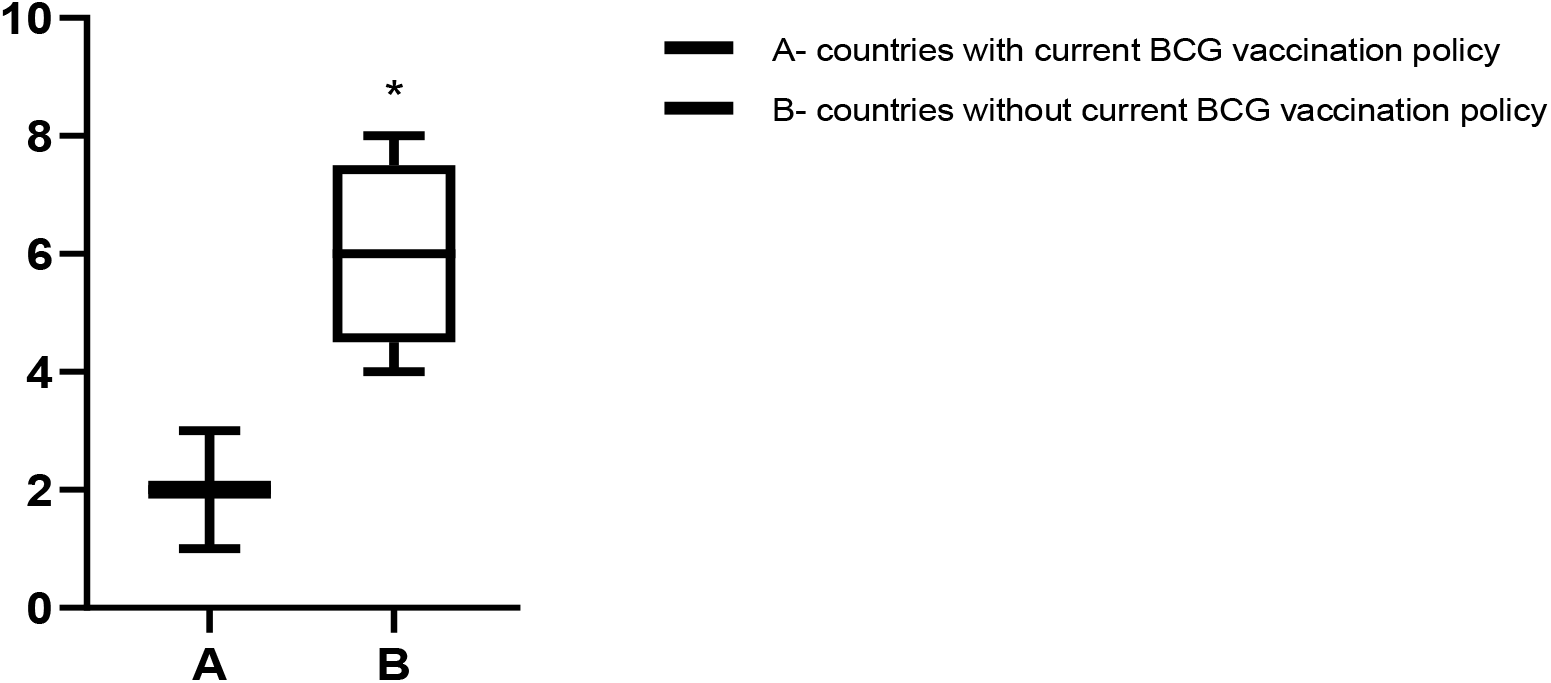
Percentage of death among the population infected with SARS-CoV-2 in **A**- countries with a current universal BCG vaccination policy (Nigeria, Ghana and South Africa) compared to **B**- countries without a current universal BCG vaccination policy (Italy, Spain, Sweden, UK and USA). Rank plot using Mann Whitney non-parametric test shows a significant (p < 0.05) difference in death rates in these two groups of countries.

**Fig. 11.**
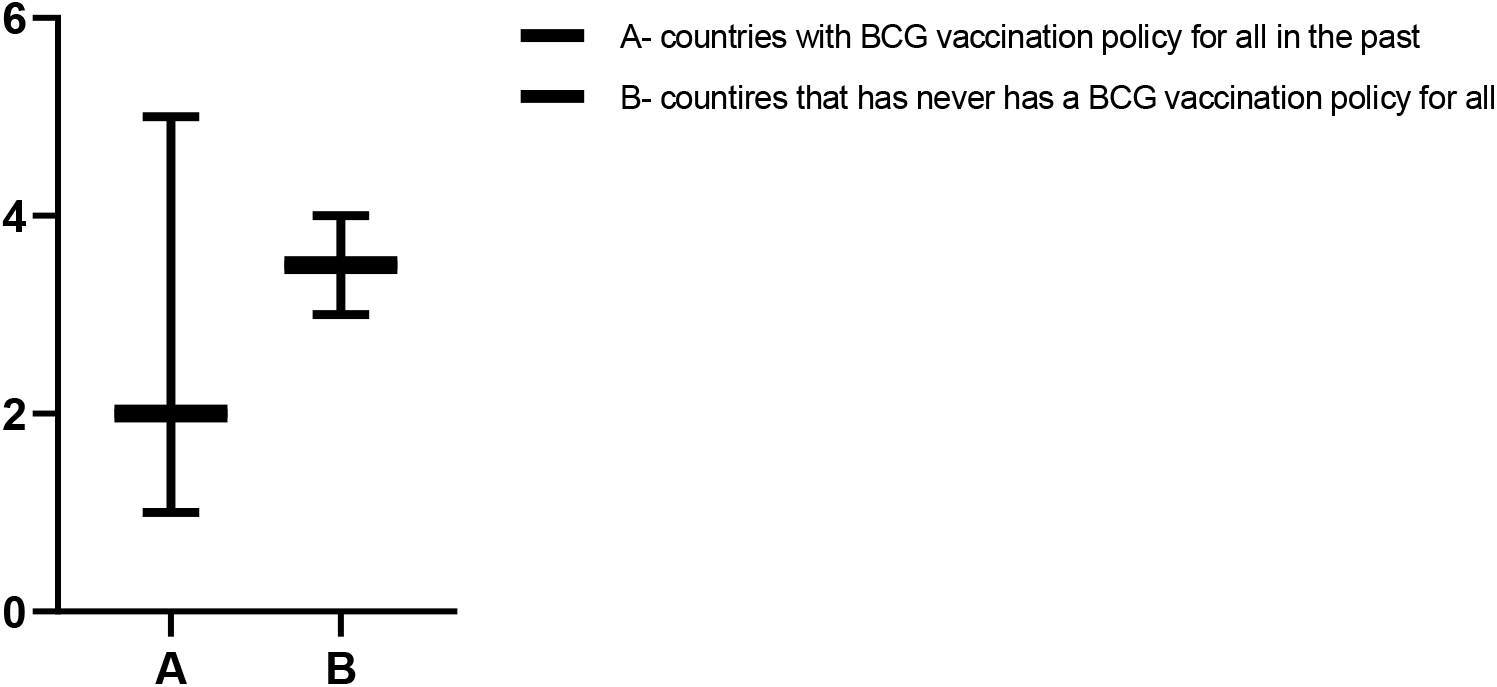
Percentage of the total population infected with SARS-CoV-2 in **A**- countries that had a universal BCG vaccination policy in the past (Spain, Sweden and UK) compared to **B**- countries that has never had a universal BCG vaccination policy (Italy and USA). Rank plot using Mann Whitney non-parametric test shows that there is no significant (p > 0.05) difference in infection rates in these two groups of countries

**Fig. 12:**
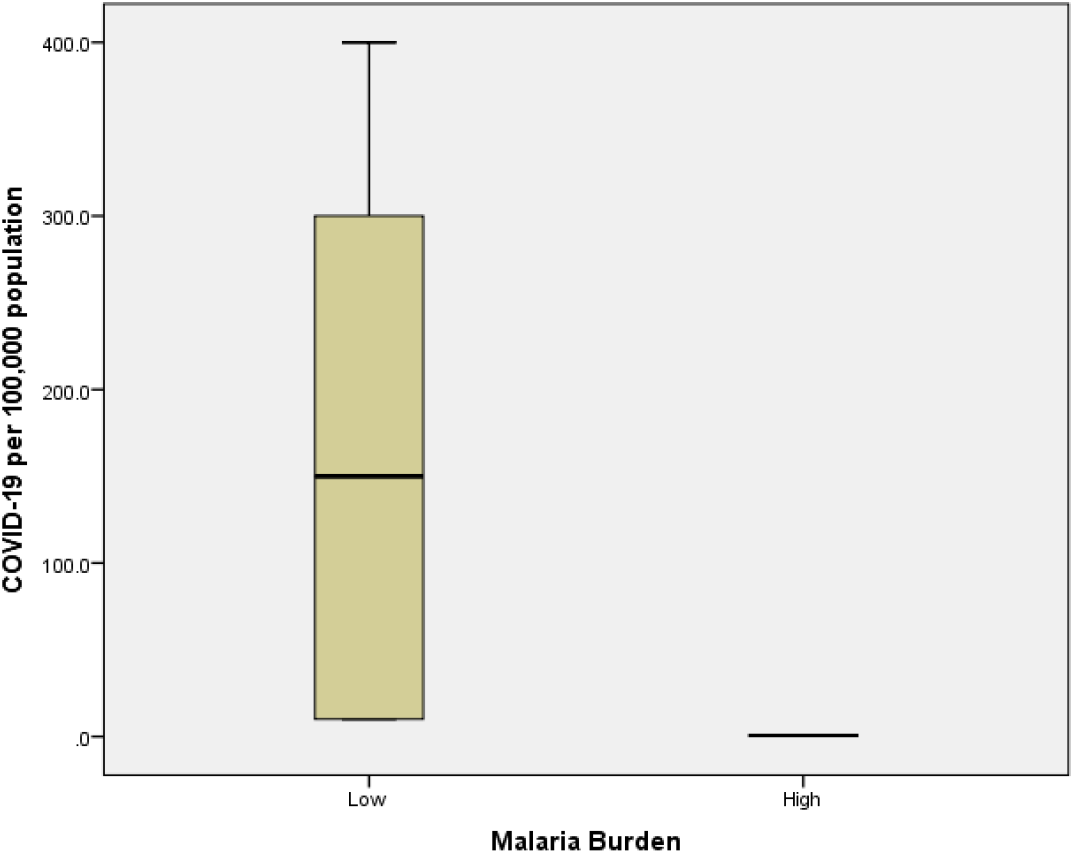
Cases of COVID-19 per 100,000 populations for countries with low malaria burden (USA, UK, Italy, Spain, Sweden and South Africa) and high malaria burden (Nigeria and Ghana). (Mann-Whitney = 0.000; p<0.05)

**Fig. 12:**
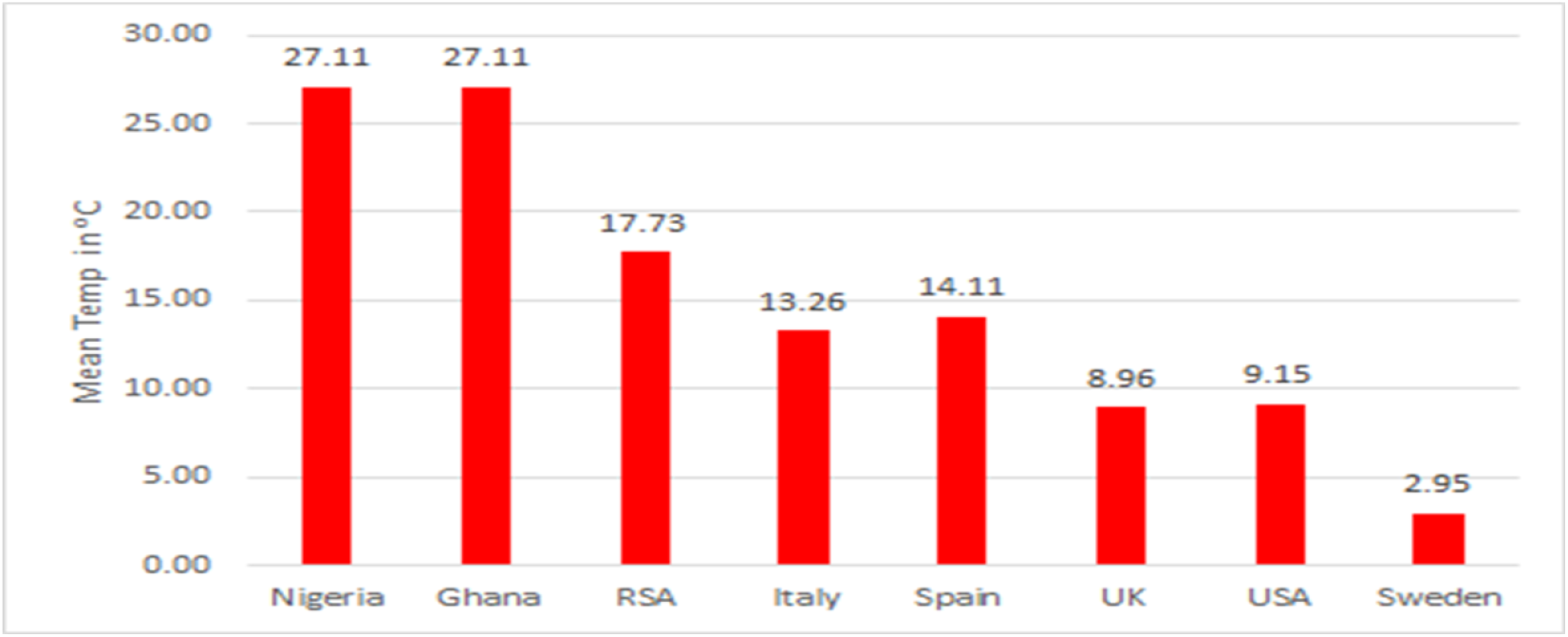
Mean Temperature for the Selected Countries (1962-2012)

### 3.4 Malaria endemicity and occurrence of COVID-19 infection

The results of the assessment of the relationship between malaria endemicity in sub-Saharan Africa and occurrence of COVID-19 are presented in Table 10 and Figure 12. The result shows there is a statistically significant (p<0.05) difference in the occurrence of COVID-19 cases between countries with high malaria endemicity compared to countries with low malaria endemicity. Countries with high malaria burden had significantly (p<0.05) lower cases of COVID-19 than those with low malaria burden. Spearman’s rank correlation was also strong. As the number of malaria cases increased, the number of COVID-19 cases decreased (r = -0.08).

**Table 10:**
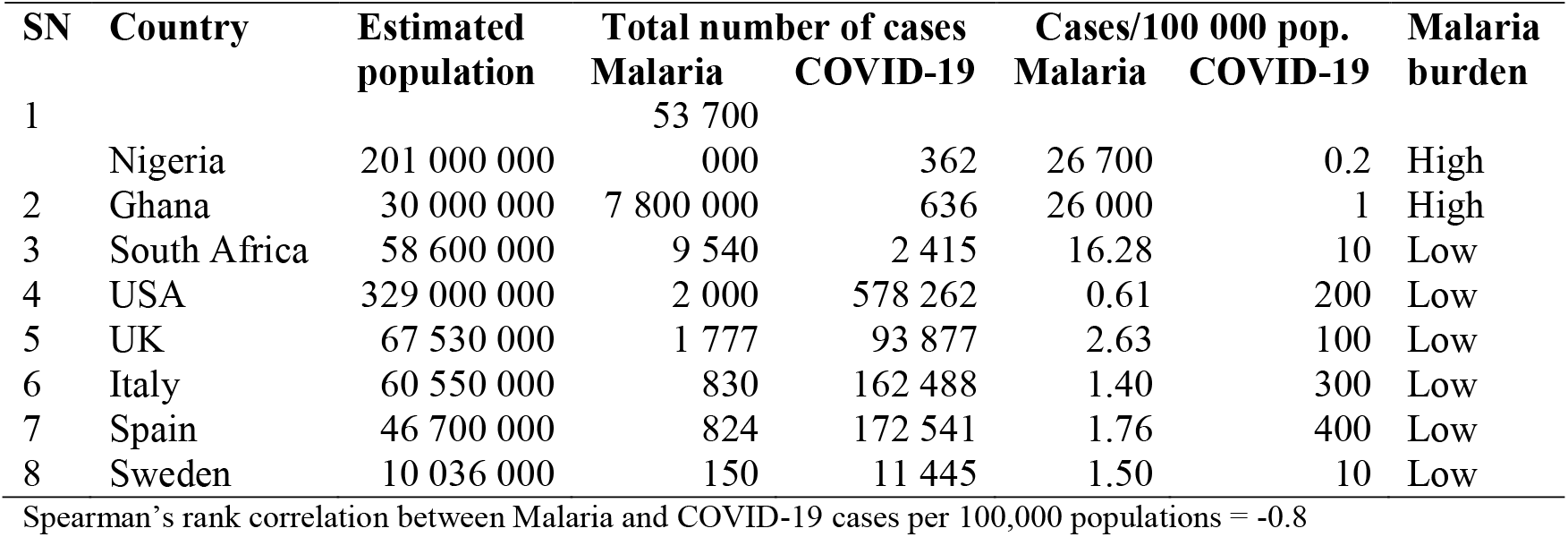
Estimated population, malaria and COVID-19 cases for the study countries

### 3.5 Climatic condition depicted by Spatial and temporal temperature pattern and COVID-19 infection

The influence of climatic condition depicted by temperature pattern on the COVID-19 infection was examined among the study countries. Figure 12 shows the computed spatial long-term mean temperature and Figure 13 shows the temporal mean monthly temperature for the selected countries. Nigeria and Ghana which are tropical countries have the highest long-term mean annual temperature with 27°C, followed by South Africa (17.7°C), Spain (14.11), Italy (13.26), USA (9.15), UK (8.96) and Sweden (2.95).

**Fig. 13:**
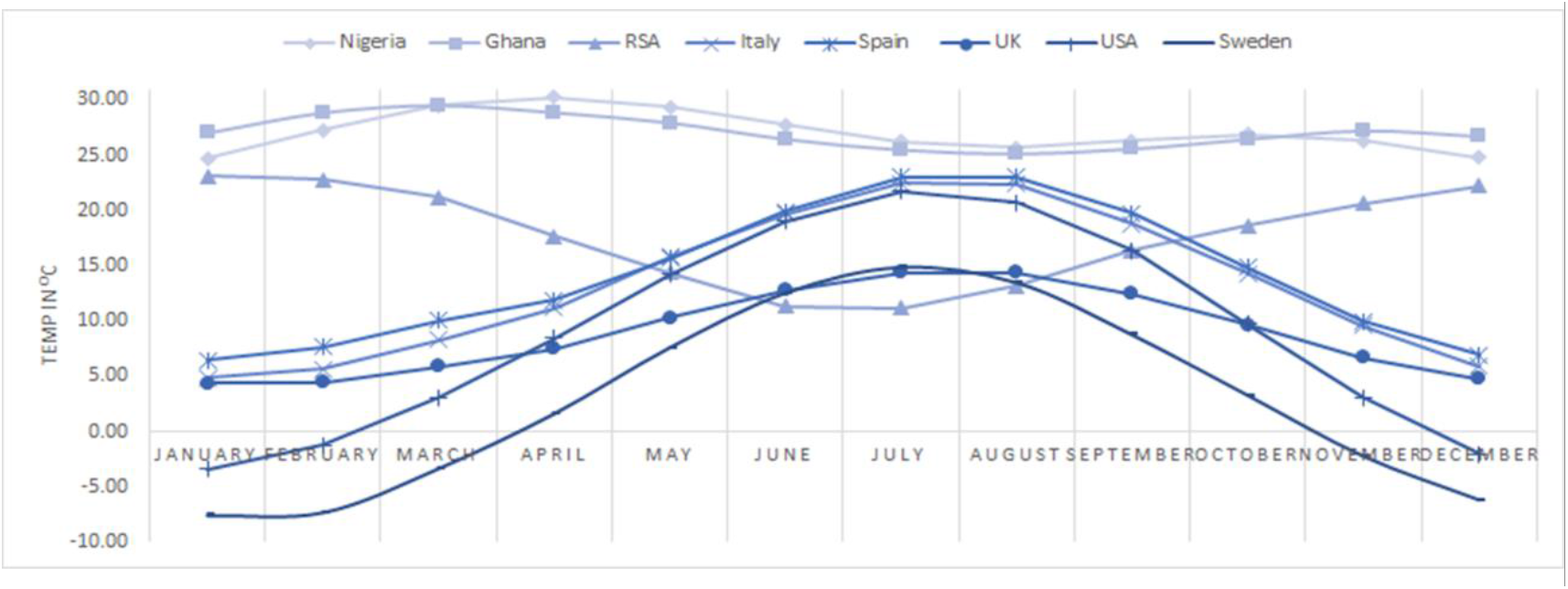
Long Term Mean Monthly Temperature for the selected countries (1962-2012)

In terms of established seasonal variation, Nigeria and Ghana have no markedly different summer and winter. Although they do experience some oscillation of dry and wet seasons, All the countries (USA, Italy, Spain and UK) of the northern mid-latitudes experience well-defined summer and winter months. South Africa on the other hand have winter and summer months (though not as pronounced as the countries of the north) which occur in opposite direction to the boreal countries (Figure 13).

#### 3.5.1 Analysis of relationship between Temperature and COVIVD-19 cases

Figure 14 is a plot of the long-term mean temperature with the COVID-19 infection cases. A casual observation suggests that countries with low mean annual temperature also correspond to those with high COVID-19 cases.

**Fig. 14:**
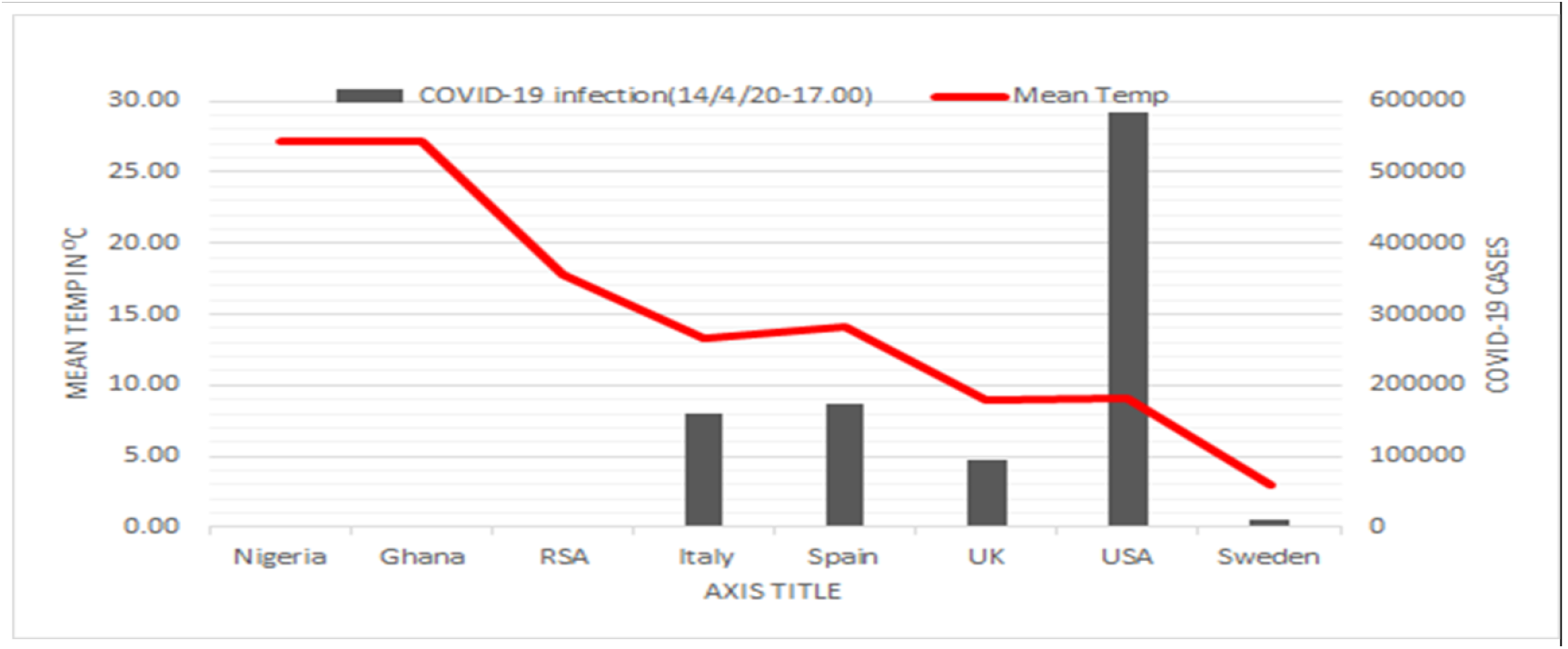
Covid-19 Cases and Average Temperature

The Spearman’s rank correlation shows a strong negative correlation between temperature and COVID-19 infection and death both having -0.595. The regression model suggests that long term mean annual temperature accounts for about 14.8% of the variances in the occurrence of COVID-19 among the countries.

Result of findings on healthcare approaches to management of COVID-19 in some African countries compared to the Europe and USA indicates a clear distinction in approach to management of the disease. Across the developed world, the preferred treatment approach has been to test, voluntary isolation at home, hospitalise for observation and treat, however, in Sub-Saharan Africa, the approach is basically test, mandatory isolation and treat (Table 11).

**Table 11:**
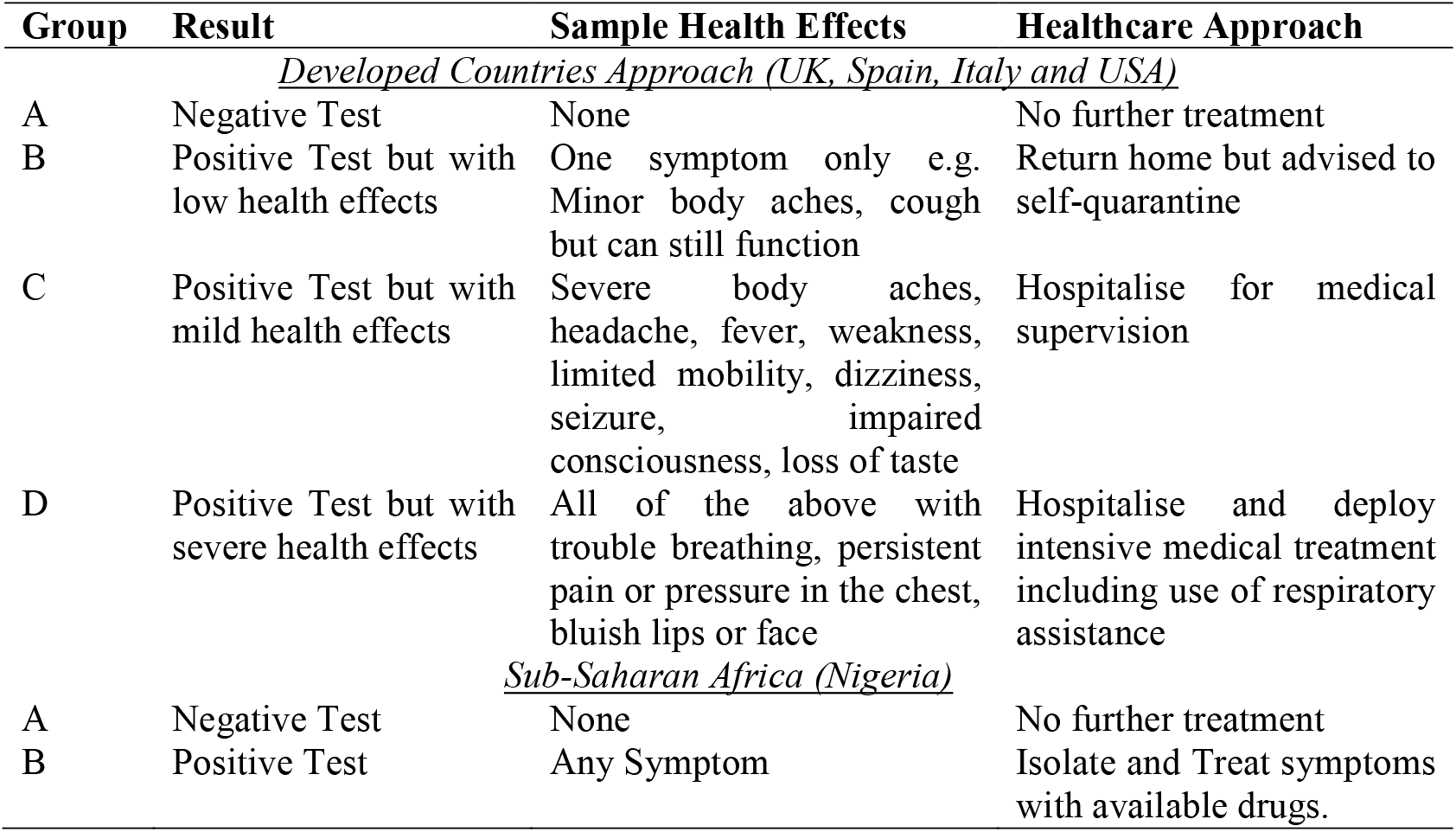
Healthcare Approaches for Management of COVID-19 in Developed Countries and sub-Saharan Africa.

## 4.0 Discussion

The outcome of the analysis of COVID-19 data for the confirmed cases of infection and recorded deaths from the index case reporting to forty-five days study period indicates a clear cut division between the different countries being assessed. In particular, countries such as Nigeria and Ghana in sub-Saharan Africa were found to have consistently different pattern of confirmed cases of COVID-19 infection and recorded death cases compared to countries with more robust and well-resourced healthcare systems in Europe (Italy, Spain and UK) and USA. During the study period, the number of confirmed cases of infection and associated deaths in sub-Saharan Africa were found to be very low (confirmed cases – range from 318 to 408; Deaths - 6 to 10) compared to the exceeding high values (confirmed cases – range from 25150 to 189967; Death – 4064 to 12428) in the developed countries with better healthcare systems.

The wide variation in reported numbers of COVID-19 infection and deaths raises a lot of concern on what might be going on with respect to epidemiology, data collection and management of the disease in the countries assessed. According to Vaughan (2020), experts still do not know why so few cases of the new coronavirus have been reported in Africa, despite China – where the virus originated – being the continent’s top trading partner and the continent having a population of 1.3 billion people. The consensus within the scientific community based on views and opinion being expressed in many online platforms is that testing for COVID-19 among the sub-Saharan African countries is generally very low, and this would have a major influence on the number of reported confirmed cases of the infection in sub-Saharan Africa. According to available data accessed on the 17^th^ of April 2020, the volume of COVID-19 tests in sub-Saharan Africa ranges from 5000 (Nigeria) to 50719 (Ghana) compared to 3.4 million in USA, 1.1 million in Italy, 930230 in Spain and 417649 in UK (www.worldodometer.com). So it is quite possible that if more testing were carried out in these countries, the number of confirmed cases will most likely rise significantly. However, given the current level of awareness of COVID-19 on the African continent, the lack of coronavirus-linked death outbreaks of the scale in Ecuador, USA, Italy, Spain or UK suggest either there is a lag in the outbreak of the disease in Africa or other factors may be responsible for the low confirmed cases of the infection and death numbers in sub-Saharan Africa compared to the overwhelming high numbers in the European countries and USA evaluated in this study.

The outcome of the evaluation to determine possibility of mutational shift that could adversely contribute to the virulence or differential incubation period of the novel corona virus (SARS-CoV-2) based on geographical location, revealed that the recovery rate from the viral infection has no correlation with the mutational attributes of the virus. The sequenced strain from Nigeria had no significant difference from other geographical regions. Ahmed *et. al*. (2020) also reported that no mutation has been observed in these identified epitopes among the 120 available SARS-CoV-2 sequences (as of 21 February 2020) and suggested that immune targeting of these epitopes may potentially offer protection against this novel virus. Our observation therefore provides a bit of relief, as regarding the lesser mutation abilities of the virus, thereby reducing the probability of longer incubation period for the epitope in Nigeria.

Results from this study have shown that there is currently a higher infection rate of SARS-CoV-2 and mortality from COVID-19 in developed countries without a current universal BCG vaccination policy compared to African countries with a current universal vaccination policy. Miller *et al*. (2020) have also reported reduced morbidity and mortality in countries with a universal BCG vaccination policy compared to countries without a policy. They indicated that the earlier the country implemented a BCG vaccination policy, the lower the number of deaths per million inhabitants. The reduced infection rates of SARS-CoV-2 and death from COVID-19 currently reported in African countries may not be a direct impact of the implementation of a universal BCG vaccination policy alone. Other factors such as climatic conditions, lifestyle and incidence of other diseases such as malaria could be responsible for the reduced incidence of COVID-19 in these countries.

The evaluation of malaria endemicity with occurrence of COVID-19 revealed that countries with high malaria burden had surprisingly low COVID-19 incidence than those with low malaria burden. This result does not infer causation, but a possible explanation could be environmental. The countries most affected are in the temperate region and viral respiratory infections usually occur more frequently during winter and fall seasons; and the COVID-19 outbreak became more widespread during the flu season in Europe and USA (Wang *et al*., 2020). For the tropical countries, flu occurs more frequently during the more humid time of the year which is the rainy reason, predominantly between June and September-October and so may explain the low infection (Haines *et al*., 2006; Lin *et al*., 2006). As a result, several opinions have been expressed on different online platforms of a possible link to the use of Chloroquine for malaria treatment. However, in many African countries, Chloroquine use has long been changed to Artemisinin Combination Therapies (ACTs) as drug of choice for malaria treatment over a decade ago due to drug resistance. Therefore, the observation of occurrence of low confirmed cases of COVID-19 with high malaria burden associated with Chloroquine use cannot be fully substantiated. It is therefore highly possible that there are other existing factors which may be responsible for the observation of high malaria endemicity with low occurrence of COVID-19 cases in this study.

Although several factors have been reported to aid the dispersal and transmission of COVID- 19 across the globe, temperature remains a strong factor of the transmission efficiency (Chan *et al*., 2011; Yip *et al*., 2017). The outcome of our study suggests that COVID-19, like the flu disease category, thrive better in low temperatures. This result is consistent with Sajadi *et al*. (2020) who reported distribution of significant community outbreaks along restricted latitude, temperature and humidity which are consistent with the behavior of seasonal respiratory virus. Yao *et al*. (2020) however submit that there was no relationship between temperature (and radiation) and COVID-19 cases in the cities of China. There is therefore the need for more studies to determine how seasonal changes in temperature (summer vs. winter) will affect the transmission of COVID-19 especially as the boreal summer approaches and the southern temperate countries prepare for winter.

Current guidance from the World Health Organisation (WHO) is that there is no known cure for this virus with 100% certainty however, research for vaccinations and cure continues fervently. In the interim, official guidance from global public health authorities is to test, test and test as many individuals of the population, practise social distancing, washing of hands with soap, use of hand sanitizers and then to proceed with caution when adopting any of the speculative treatment options. This very loose guidance has led to differential healthcare approaches to management of COVID-19 in different countries across the world. The outcome of our findings has revealed that fundamental differences exist between the healthcare systems of sub-Saharan Africa and those of Europe and America. In Europe and America, the preferred treatment approach has been to test, voluntary isolation at home, hospitalise for observation and treat, however, in sub-Saharan Africa, the approach is basically test, mandatory isolation and treat (see Table 11). So while the developed countries are concentrating on testing, the sub-Saharan African countries seem to be concentrating more on treating the diseases with some of the speculative re-purposed drugs.

Although there seems to be no basis for comparison in terms of resources and organisation between the healthcare systems in Europe and USA with that of sub-Saharan Africa, the outcomes with regards to number of recorded deaths or recovery rate of COVID-19 patients is staggering and very baffling. While a more resourced healthcare system in Europe and America has an outcome of 4064 (USA), 1789 (UK), 8464 (Spain) and 12428 (Italy) deaths after 44 days of reporting the 1^st^ index case, a country like Nigeria recorded 10 deaths after 44 days (www.worldodometer.com). So it does appear that something must be amiss because the numbers simply do not follow basic logic. It therefore seems that there may be a need to review the current healthcare approaches or guidance the mentioned developed countries have adopted to managing the COVID-19 illness. The outcomes in terms of the substantial huge number of recorded deaths seem to indicate a weakness in the healthcare approach. It may just be time to learn a lesson or two from the approach which the less developed countries have adopted to manage the disease in this time of emergency.

In Europe and America, the healthcare system is largely centrally controlled; public sector based and with rampant initiation of professional consequences if a wrong call is made. Also, there is controlled access to many drugs. However, in sub-Saharan Africa countries like Nigeria, the healthcare system is predominantly private, with lesser occurrence of initiation of professional consequences. The inability or difficulty of the healthcare workers in the more developed countries to adapt rapidly and explore the use of speculative drugs during this emergency maybe attributable to the stringent medical guidance, professional consequences and exposure to lawsuits associated with medical care in the countries. However, the differential outcomes from the two approaches in the management of this pandemic between the selected countries is indicative that in the enactment of emergency legislations for disease outbreaks, the congress or legislative bodies need to consider inclusion of clauses which are aimed at shielding the healthcare professionals of potential litigations that may arise from some decisions taking in their attempts to save lives during this type of novel disease outbreaks.

Apart from healthcare approach to management of the disease, a curious look at the healthcare systems in the different countries points to major obvious differences. Aside from the substantial differences in funding and manpower availability, there is almost unfettered access to most types of prescription drugs. Therefore, it is very possible that a large number of COVID-19 positives in Africa would have reached out to the pharmacist for different treatment sessions before eventually going for COVID-19 test. So could this early treatment occasioned by easy access to prescription drugs be a factor in the lowering death numbers being recorded in countries like Nigeria and Ghana? There is little doubt that in an emergency situation like the COVID-19 pandemic, the somewhat informal nature of healthcare practice in sub-Saharan Africa probably gives the healthcare professionals more room to operate by prescribing without formal guidance, certain drugs which have been reported in several media as being effective for the treatment of COVID-19, without significant fear of professional consequences and litigations that are quite rampant in the health administration of the more developed countries. There is also little doubt that lessons learnt from the Ebola incident in sub-Saharan Africa may have also contributed to the low numbers of COVID-19 confirmed cases and deaths. Communities that experienced Ebola in West Africa in 2014-2016 seem to have learnt a painful lesson on how to cope with a deadly new infection with important information on strategies to address novel disease threats more generally (The World Bank, 2020). Nigeria for example, announced on Friday 28 January 2020 the first index case coronavirus disease COVID-19 in sub-Saharan Africa’s region and the confirmation led to the immediate activation of the country’s National Coronavirus Emergency Operation Centre riding on facilities put in place during the Ebola crisis (Nature Medicine, 2020). There is no doubt that when another epidemic arises again, the experiences of the COVID-19 pandemic will make a lot countries to act differently and possible swiftly, which might affect the overall disease burden.

## Data Availability

Data used for the study are available at online database platforms

http://coronavirus.jhu.edu/

http://www.worldometers.info/

## Limitation of the study

As the covid-19 pandemic is still unfolding, this paper has examined several factors in spatial differences associated with the disease over a 46 period of observation. The patterns and trends are still evolving and by the time the paper is published, significant changes could have occurred. The wide variation in the number of COVID-19 testing being carried out by the different countries will also have a major impact on the actual number of reported confirmed cases of infection. The number of deaths associated with COVID-19 during the period of study is however likely to follow the same trend as reported. A few selected countries have been chosen for evaluation and this may limit generalizability of findings. The findings however cuts across continents and this will provide scientific basis for detailed and more targeted research.

## Conclusion

This study has revealed compelling spatial differences in the incidence and deaths from COVID-19 in selected countries in sub-Saharan Africa compared to Europe and USA over a 46-day observation period. The major factors attributed to the wide variation in the outcome of the COVID-19 disease burden in the countries examined are BCG vaccination policy, malaria endemicity, climatic condition (temperature) and differential healthcare approaches to management of the disease. The need to consider inclusion of clauses in national emergency legislations aimed at protecting healthcare professionals from lawsuits that can arise from decisions taking in their attempts to save lives during this type of novel disease outbreaks was proffered. Further research is needed to confirm whether the spatial differences in incidence and deaths are sustained when the pandemic is resolved globally.

## Ethical approval

This study did not involve human or animal experimentation. All data sources are cited and acknowledged throughout the manuscript.

## Acknowledgement

The authors acknowledge the opportunity to serve as volunteers of the UNILAG Consult COVID-19 Advisory Group (UCCAG).

## Authors’ contribution statement

The authors have contributed equally to the publication.

## Websites

▪ www.aljazeera.com/news/2020/04/police-collect-800-bodies-ecuador-virus-epicenter-200413082635865.html (Accessed online: 14/04/2020 at 9.57 Nigerian Time).
▪ www.who.int/emergencies/diseases/novel-coronavirus-2019/events-as-they-happen (Accessed online: 14/04/2020 at 10.32am Nigerian Time)
▪ https://www.cdc.gov/malaria/resources/pdf/fsp/cdc_malaria_domestic_unit.pdf
▪ https://stat.world/biportal/?lang=en&solution=Climate+Statistics&allsol=1
▪ https://nairametrics.com/2020/04/08/analysis-nigerias-likely-cost-per-treatment-for-covid-19/ Accessed online: 14/04/2020 @ 4:47pm Nigerian Time
▪ https://coronavirus.jhu.edu/
▪ https://www.worldometers.info/
▪ https://github.com/acegid/CoV_Sequences/
▪ https://www.bcgatlas.org/
▪ https://ecdc.europa.eu/
▪ https://stat.world/biportal/?allsol=1
▪ https://www.healthdata.org/

## References

1. Ahmed, S.F., Quadeer, A.A. and McKay, M.R. (2020). Preliminary identification of potential vaccine targets for the COVID-19 Coronavirus (SARS-CoV-2) based on SARS-CoV Immunological Studies. Viruses 12(3): 254. https://doi.org/10.3390/v12030254.

2. Baker, A. (2020). Few doctors, few ventilators: african countries fear they are defenseless against inevitable spread of coronavirus. www.time.com/5816299/coronavirus-africa-ventilators-doctors/

3. Chan, K.H., Malik Peiris, J.S., Lam, S.Y., Poon, L.L.M., Yuen, K.Y. and Seto, W.H. (2011). The Effects of temperature and relative humidity on the viability of the SARS Coronavirus. Adv Virol 1687–8639. https://doi.org/10.1155/2011/734690

4. Cucinotta, D. and Vanelli, M. (2020). WHO declares COVID-19 a pandemic. Acta Biomed 91(1): 157–160. doi: 10.23750/abm.v91i1.9397.

5. Ebenso, B. and Otu, A. (2020). Can Nigeria contain the COVID-19 outbreak using lessons from recent epidemics? Lancet Glob Health 30101-7. DOI: https://doi.org/10.1016/S2214-109X(20)30101-7.

6. European Centre for Disease Prevention and Control (ECDC) (2019). Malaria. In: ECDC. Annual epidemiological report for 2017. ECDC, Stockholm. Retrieved from ecdc.europa.eu.

7. Gilbert, M. Pullano, G., Pinotti, F., Valdano, E., Poletto, C., Boelle, P., D’Ortenzio, E., Yazdanpanah, Y., Eholie, S.P., Altmann, M., Gutierrez, B., Kraemer, M.U.G. and Colizza, V. (2020). Preparedness and vulnerability of African countries against importations of COVID-19: a modelling study. The Lancet, 395(10227): 871-7. DOI: https://doi.org/10.1016/S0140-6736(20)30411-6.

8. Haines, A., Kovats, R.S., Campbell-Lendrum, D. and Corvalan. C. (2006). Climate change and human health: impacts, vulnerability, and mitigation. Lancet, 367: 2101–09.

9. Hammer, O., Harper, D.A.T. and Ryan, P.D. (2001). PAST 3.2. Paleontological statistics software package for education and data analysis. Palaeontol Electron 4(1): 9pp.

10. Lin, K., Yee-Tak Fong, D., Zhu, B. and Karlberg, J. (2006). Environmental factors on the SARS epidemic: air temperature, passage of time and multiplicative effect of hospital infection. Epidemiol Infect 134(2): 223–30.

11. Martinez-Alvarez, M., Jarde, A., Usuf, E., Brotherton, H., Bittaye, M., Samateh, A.L., Antonio, M., Vives-Tomas, J., D’Alessandro, U. and Roca, A. (2020). COVID-19 pandemic in West Africa. Lancet Glob Health Online First. DOI: https://doi.org/10.1016/S2214-109X(20)30123-6.

12. Miller, A., Reandelar, M.J., Fasciglione, K., Roumenova, V., Li, Y. and Otazu, G.H. (2020). Correlation between universal BCG vaccination policy and reduced morbidity and mortality for COVID-19: an epidemiological study. medRxiv preprint doi: https://doi.org/10.1101/2020.03.24.20042937

13. Nature Medicine (2020). Nigeria responds to COVID-19; first case detected in sub-Saharan Africa. https://www.nature.com/articles/d41591-020-00004-2

14. Ogbeibu, A. E. (2014). Biostatistics – A Practical Approach to Research and Data Handling. Mindex Press, Benin City, Nigeria. 285pp.

15. Sajadi, M.M, Habibzadeh, P, Vintzileos, A, Shokouhi, S, Miralles-Wilhelm, F, Amoroso, A (2020). Temperature, humidity, and latitude analysis to predict potential spread and seasonality for COVID-19. https://ssrn.com/abstract=3550308.

16. Stamatakis, A. (2006). RAxML-VI-HPC: Maximum likelihood-based phylogenetic analyses with thousands of taxa and mixed models. Bioinformatics 22, 2688–2690.

17. The World Bank (2020). In the face of Coronavirus, African countries apply lessons from Ebola response. https://www.worldbank.org/en/news/feature/2020/04/03/.

18. United Nations Economic Commission for Africa (ECA) (2020). COVID-19 in Africa: Protecting Lives and Economies. ECA Printing Publishing Unit, 38pp.

19. Vaughan, A. (2020). We don’t know why so few COVID-19 cases have been reported in Africa. https://www.newscientist.com/article/2236760.

20. Wang, J., Tang, K., Feng, K. and Lv, W. (2020). High temperature and high humidity reduce the transmission of COVID-19. http://dx.doi.org/10.2139/ssrn.3551767.

21. World Health Organisation (2019). World Malaria Report. WHO, Geneva: Licence: CC BY-NC-SA 3.0 IGO.

22. World Health Organisation (2020). Coronavirus disease 2019 (COVID-19): Situation report – 88. WHO, Geneva: 12pp.

23. Yao Y. et al.. (2020). No Association of COVID-19 transmission with temperature or UV radiation in Chinese cities. Eur Respir J In Press. https://doi.org/10.1183/13993003.00517-2020.

24. Yip, C., Chang, W.L., Yeung, K.H., and Yu, I.T. (2007). Possible meteorological influence on the severe acute respiratory syndrome (SARS) community outbreak at Amoy Gardens, Hong Kong. JEH 70(3):39–46.

